# WALRUS: Robust and Transferable Deep Learning for Water and Lipid Removal in ^1^H-MRSI

**DOI:** 10.64898/2026.11.09.26366974

**Authors:** Hauke Fischer, Stanislav Motyka, Paul Weiser, Bernhard Strasser, Lukas Hingerl, Korbinian Eckstein, Korawig Chaknam, Aaron P. Osburg, Anton Altmeyer, Bernardo Campilho, Ekaterina Sazonova, Yan Li, Jing Liu, Barbara Dymerska, Zeinab Eftekhari, Thomas B Shaw, Markus Barth, Gilbert Hangel, Sumit Kaushik, Lukas Haider, Wolfgang Bogner

## Abstract

Proton magnetic resonance spectroscopic imaging (^1^H-MRSI) enables non-invasive mapping of brain metabolism, but reliable quantification is challenged by residual water and extracranial lipid contamination. Recent work demonstrated that a deep learning model trained on simulated metabolite spectra combined with *in vivo*-derived water and lipid signals can outperform conventional nuisance-removal methods. However, this approach relied on heuristic simulation parameter distributions and a subject-specific L2-derived lipid estimate as an auxiliary network input, requiring subject-specific lipid masks and L2 processing during inference. Here, we introduce WALRUS (Water And Lipid Removal Using Spectral Learning), a fitting-consistent and empirically calibrated framework developed through a systematic evaluation of simulation realism, architectural priors, and network optimization. WALRUS uses the same metabolite basis as downstream spectral fitting and calibrates deliberately broadened simulation distributions from *in vivo* measurements. Controlled 7 T experiments demonstrate that empirical simulator calibration substantially improves nuisance removal and metabolite quantification compared with heuristic training distributions. With the optimized simulator, a capacity-matched comparison reveals no clear additional benefit from the L2-derived prior, allowing its removal and substantially simplifying inference. Further improvements are achieved with the final higher-capacity U-Net, while on-the-fly simulation improves generalization compared with training on a fixed synthetic dataset. The same framework was subsequently recalibrated and retrained at 3 T, where WALRUS outperformed conventional HLSVD+L2 processing on held-out data. Together, WALRUS combines these advances into an optimized framework that improves nuisance removal and downstream metabolite fitting, requires no subject-specific lipid segmentation or L2-derived auxiliary input during inference, and can be systematically adapted to different acquisition settings.

## 1 Introduction

Proton magnetic resonance spectroscopic imaging (^1^H-MRSI) enables non-invasive mapping of brain metabolism and provides information complementary to conventional structural MRI. It has been applied to a broad range of neurological and oncological conditions, including the investigation of neurochemical alterations, brain-tumor characterization, and treatment response [1–4].

Reliable quantification, however, remains challenging because metabolite signals are several orders of magnitude weaker than the dominant water signal and can additionally be contaminated by extracranial lipids [5, 6]. These nuisance signals can distort the spectral baseline, overlap with metabolite resonances, and substantially impair subsequent spectral fitting and metabolite mapping.

Lipid contamination is particularly challenging in ^1^H-MRSI acquisitions employing whole-slice or slab excitation and short or negligible echo times, where strong extracranial lipid signals can contaminate brain spectra through the spatial point-spread function [7, 8].

A wide range of strategies have been developed to suppress water and lipid contamination. These include acquisition-based approaches using dedicated RF pulses or sequence designs [9–16], hardware-based methods for suppressing extracranial lipid signal [17, 18], and post-processing techniques exploiting spatial or spectral priors [19–29].

However, acquisition-based suppression may remain incomplete due to spatial *B*_0_ and 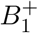 inhomo-geneities and imperfect extracranial lipid suppression [30]. Additional post-processing is therefore often required. For residual-water removal, Hankel–Lanczos singular value decomposition (HLSVD) is a widely used spectral decomposition method [27], whereas lipid contamination can be reduced using spatial and spectral prior-based approaches, including L2 regularization and related subspace methods [22, 24, 31]. Such methods can be effective, but their performance depends on assumptions about the nuisance-signal subspace and on subject-specific choices such as anatomical mask definition and regularization strength. When these assumptions are not fully satisfied, residual nuisance signal may remain or metabolite signal may be attenuated. Therefore the development of more robust and less subject-dependent approaches are needed.

More recently, WALINET (WAter and LIpid neural NETwork) demonstrated that supervised deep learning can provide an effective alternative to conventional water- and lipid-removal methods [32]. WALINET combined simulated metabolite spectra with water and lipid signals extracted from measured *in vivo* MRSI data and incorporated a subject-specific L2-derived lipid projection as an auxiliary network input. This hybrid approach learned to estimate nuisance signals directly, achieving lower residual errors in simulated data and improved SNR and CRLBs *in vivo* compared with conventional HLSVD+L2 processing. A subsequent extension, WALINET+, applied the same general concept to water-unsuppressed MRSI and additionally targeted gradient-induced water sidebands [33].

Despite these advances, several fundamental questions regarding the design of simulation-based nuisance-removal frameworks remain open. Previous work has demonstrated that simulation-based deep learning can outperform conventional methods, using broad heuristic simulation distributions and auxiliary inputs derived from classical nuisance-removal techniques. However, it remains unclear to what extent more realistic simulations can further improve *in vivo* performance, whether the L2-derived auxiliary input provides additional benefits, and how much further improvement can be achieved through network optimization.

Here, we introduce WALRUS (Water And Lipid Removal Using Spectral Learning), a fitting-consistent and empirically calibrated framework designed to address these questions. A central design principle is to build the simulator around the same spectral model used for downstream metabolite quantification.

This ensures consistency between simulation and fitting, minimizes additional modeling dependencies, and enables simulation parameters to be calibrated from *in vivo* spectral fitting results.

To systematically investigate the factors governing nuisance-removal performance, we follow a three-step development strategy (Fig. 1). First, rather than assuming the necessity of additional architectural priors, we investigate how much performance can be improved through simulator optimization alone, using U-Nets with matched network capacity and training conditions. Second, after optimizing the simulator, we assess whether an additional L2-derived prior provides further benefit by comparing capacity-matched U-Net and WALINET-style Y-Net architectures. Finally, we investigate whether the resulting framework can be further improved using higher-capacity networks, and evaluate the role of on-the-fly simulation in supporting their training.

**Figure 1:**
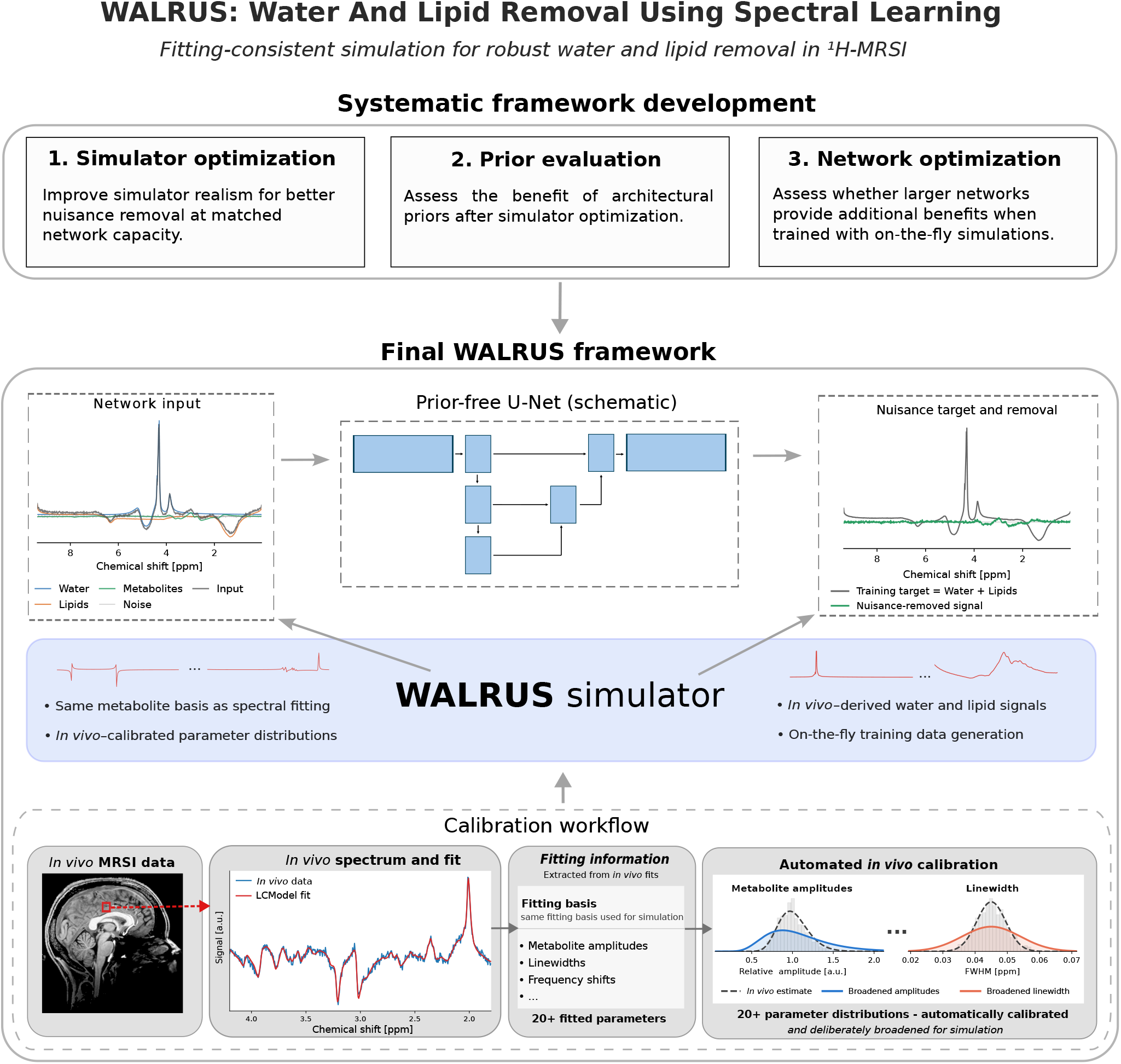
Overview of the WALRUS development strategy and final framework. Systematic model development. The development follows three sequential steps: (1) improving simulation realism to enhance nuisance removal at matched network capacity, (2) evaluating the additional benefit of an L2-derived auxiliary input as an architectural prior after simulator optimization, and (3) investigating whether larger networks provide further improvements when trained with on-the-fly simulations. **Final WALRUS framework**. *In vivo* MRSI data are spectrally fitted to derive field-strength-specific distributions of metabolite amplitudes and spectral parameters. These distributions are robustly calibrated and deliberately broadened to define the sampling distributions of the WALRUS simulator, which uses the same metabolite basis as spectral fitting together with *in vivo*-derived residual-water and lipid signals. Synthetic spectra comprising metabolites, residual water, lipids, and noise are generated on the fly during training. The network is trained to predict the combined nuisance signal consisting of water and lipids, which is subsequently subtracted from the input spectrum to obtain the nuisance-removed signal.

Beyond the systematic development at 7 T, we investigate the transferability of WALRUS to 3 T by applying the same framework with field-strength-specific spectral fitting bases and empirically recalibrated simulation distributions. This allows us to assess whether the resulting approach can be systematically adapted to substantially different acquisition settings while maintaining competitive performance against conventional nuisance-removal methods.

## 2 Materials and Methods

Written informed consent was obtained from all participants that underwent MRSI scans for the acquisition of the *in vivo* datasets used in this study.

The Vienna acquisitions were approved by the Ethics Committee of the Medical University of Vienna (approval number EK 1767/2022) and was conducted in accordance with the Declaration of Helsinki. The Brisbane acquisitions were approved by the University of Queensland Human Research Ethics

Committee (HREC) under the Biomarkers of Long Surviving MND (BeLong) study (2021/HE000975). The London acquisitions were approved by the UCL Life and Medical Sciences Research Ethics Committee under the METAHEAD study (Project ID: 0584).

The UCSF acquisitions were approved by the University of California, San Francisco Institutional Review Board (UCSF IRB) (IRB Number: 23-38347).

### 2.1 Data and preprocessing

^1^H-MRSI data were acquired using three-dimensional concentric-ring-trajectory free-induction-decay MRSI (3D-CRT-FID-MRSI) [34, 35] at 3 T and 7 T, alongside anatomical T_1_-weighted imaging for voxel positioning and tissue segmentation. At 7 T, data were acquired from five healthy volunteers at each of four sites (Vienna, Brisbane, London, and UCSF).

At 3 T, data were acquired from five healthy volunteers per scanner across four scanners in Vienna and Brisbane. Multiple acquisition configurations yielded 61 MRSI datasets. Detailed acquisition parameters and the distribution of 3 T datasets across scanners and spatial resolutions are provided in Supplementary Tables S1 and S2.

At 7 T, four subjects from each of the Vienna, Brisbane, and London sites were assigned to training and one to validation (12 training and three validation subjects). The validation subjects were used to guide methodological decisions during framework development, while the held-out UCSF subjects were reserved for independent final evaluation. At 3 T, four subjects per scanner were assigned to training and one to evaluation, yielding 49 training and 12 held-out MRSI datasets across the different acquisition configurations. All simulator calibration parameters and empirical water and lipid resources were derived exclusively from the training data, with held-out test subjects excluded.

MRSI data were reconstructed offline using an established processing pipeline [36], including iMUSICAL coil combination, non-Cartesian spatial reconstruction, and B_0_ correction. The resulting complex-valued spectra were used for nuisance removal and spectral fitting.

Voxel-wise spectral fitting was performed using LCModel (v6.3.1) [37] over 1.8–4.2 ppm with field-strength-specific basis sets generated from quantum-mechanical simulations [38, 39]. The 3 T metabolite basis comprised Cr, GABA, GPC, GSH, Gln, Glu, Ins, NAA, NAAG, PCh, and PCr. At 7 T, Asp, Glc, scyllo-inositol, and Tau were additionally included. A measured macromolecular background was included in the LCModel fitting basis at both field strengths [40]. LCModel-derived metabolite amplitudes, SNR, frequency shifts, phase parameters, and linewidths were retained for WALRUS simulator calibration. The same metabolite basis functions were used in the WALRUS simulator. For evaluation, metabolite maps were generated for tNAA (NAA + NAAG), tCr (Cr + PCr), tCho (GPC + PCh), and Glx (Glu + Gln).

### 2.2 Study design

The methodological framework and experimental strategy are organized as presented in Fig. 1.

Section 2.3 introduces the fitting-consistent spectral simulator and two alternative parameterizations: heuristic distributions based on previous work [32] and empirically calibrated distributions. These provide the basis for investigating the influence of simulation realism independently of additional architectural priors, using networks with matched capacity and training conditions.

Section 2.4 describes the conventional HLSVD+L2 reference pipeline and the construction of an L2-derived auxiliary input to the Y-Net, allowing us to investigate whether classical prior information provides additional benefit once the simulator has been calibrated.

Section 2.5 describes the network architectures and training procedures, including the higher-capacity U-Net trained with on-the-fly simulations.

The controlled comparisons and evaluations and the adaptation of the framework to 3 T are detailed in Section 2.6.

### 2.3 Spectral simulation

#### 2.3.1 Fitting-consistent spectral simulator

Each simulated spectrum was constructed as the sum of metabolite, residual-water, lipid, and receivernoise components,

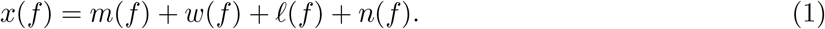

Here, *f* denotes the spectral frequency coordinate, while *m, w, l*, and *n* represent the metabolite, residual-water, lipid, and receiver-noise components, respectively. The simulator was designed to follow the parameterization used for spectral fitting as closely as possible. The metabolite component was generated from the same field-strength-specific basis sets used for LCModel fitting (Sec. 2.1). Relative metabolite amplitudes were sampled independently and a common frequency shift, zero- and first-order phase, and Voigt-type line broadening were applied to the resulting metabolite mixture. Complex receiver noise was subsequently added according to a sampled SNR.

Residual-water and lipid line shapes were derived from measured *in vivo* data rather than generated analytically. Residual-water signals estimated by HLSVD [27] from voxels within the brain mask formed subject-specific water pools. Lipid resources were obtained from spectra in the extracranial lipid mask after removal of the corresponding residual-water component. During simulation, water and lipid signals were sampled from these empirical resource pools and scaled relative to the simulated metabolite signal.

The simulator additionally supported variable acquisition lengths by truncating the individual signal components in the time domain before final spectrum assembly. The complete simulated spectrum was normalized by its maximum spectral magnitude, and the corresponding training target consisted of the combined residual-water and lipid components normalized by the same factor. Thus, the network was trained to predict the nuisance signal directly, which was subsequently subtracted from the input spectrum during inference.

Full mathematical details of the spectral model, empirical resource sampling, noise scaling, and signal normalization are provided in Supplementary Note S1.1. Representative simulated spectra at both field strengths are shown in Supplementary Fig. S3.

#### 2.3.2 Empirically calibrated distributions

Two distribution families were used to sample the simulator parameters: *Z*_+_ for positive-valued parameters, such as metabolite amplitudes, linewidths, and SNR, and *Z*_*±*_ for signed parameters, such as frequency shifts. For positive-valued parameters, an equally weighted mixture of a zero-truncated normal and a lognormal distribution was used. While the lognormal distribution provides an extended positive tail, its probability density approaches zero near the origin, potentially undersampling very small parameter values. The zero-truncated normal component addresses this limitation by maintaining a non-zero probability density near zero.

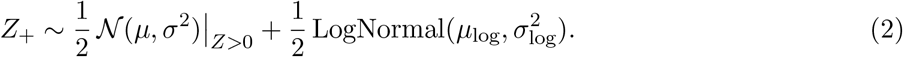

A lognormal random variable *Z* is defined by

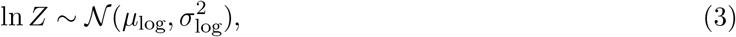

where *µ*_log_ and *σ*_log_ denote the mean and standard deviation in logarithmic space.

For signed parameters, a symmetric mixture consisting of a central normal component and positive and negative lognormal tails was used. The normal component provides coverage around the distribution center, while the lognormal components extend the sampled range toward both positive and negative extremes

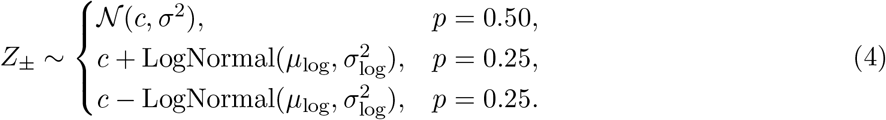

The distributions were calibrated separately for 3 T and 7 T using parameter estimates derived from *in vivo* spectral fitting. Distribution centers and widths were estimated robustly from the median and interquartile range (IQR),

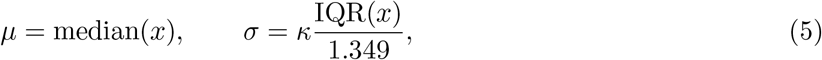

where *κ* controls the broadening of the sampled distributions beyond the observed variability. A broadening factor of *κ* = 2 was used, except for linewidth (*κ* = 1). Lognormal components were calibrated analogously in logarithmic space, while signed distributions were centered on the median and their tails calibrated from absolute deviations. Detailed calibration procedures, including metabolite amplitudes and water- and lipid-to-metabolite ratios, are provided in Supplementary Note S1.

#### 2.3.3 Heuristic distributions

As an alternative to empirical calibration, heuristic parameter distributions based on previous work [32] were implemented within the same spectral simulator. Distributions were adopted where they could be mapped to the corresponding simulator parameters. Metabolite amplitudes were sampled independently from a zero-truncated normal distribution with mean 1 and standard deviation 5. Global frequency shifts were sampled uniformly between *−*0.5 and 0.5 ppm, Voigt FWHM between 0.0135 and 0.168 ppm, and zero-order phase between 0 and 2*π*. Water-to-metabolite and lipid-to-metabolite peak amplitude ratios were sampled log-uniformly over 0.01–100 and 0.1–1000, respectively.

The empirically calibrated SNR distribution was retained instead of adopting the original WALINET distribution, because SNR was defined differently in the WALRUS simulator (see Supplementary Note S1.1). Similarly, the empirically calibrated first-order phase distribution was retained because this parameter is closely linked to acquisition- and reconstruction-specific characteristics.

### 2.4 Conventional nuisance removal and auxiliary prior

#### 2.4.1 Conventional HLSVD+L2 processing

Residual water was estimated using rank-32 HLSVD [27], reconstructing spectral components between 4.14 and 6 ppm. The same procedure was used for conventional water removal, preparation of empirical water and lipid resources, and simulator calibration.

Lipid suppression was performed using a subject-specific L2 regularization approach [32]. For each subject, spectra from the extracranial lipid mask were first water-suppressed using HLSVD and collected in a matrix *R*. The L2 regularization operator and corresponding lipid projection were defined as

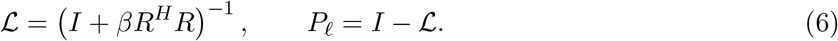

Here, 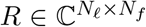 contains *N*_*l*_ water-suppressed lipid-mask spectra as rows, each with *N*_*f*_ spectral points. The matrix 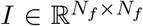 is the identity matrix, *R*^*H*^ denotes the conjugate transpose of *R*, and *β ≥* 0 is the real-valued regularization parameter. Following [32], *β* was adjusted to achieve a specified mean absolute diagonal value of *L, τ* = mean(| diag(*L*)|), hereafter referred to as the L2 trace target.

For a complex-valued row spectrum 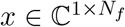, the estimated lipid component was given by 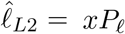 . The L2 trace targets were selected by visual inspection of representative spectra to achieve a reasonable compromise between lipid suppression and metabolite preservation (Supplementary Fig. S4). For the conventional HLSVD+L2 reference, water and lipid components were estimated independently from the original spectrum and subtracted according to

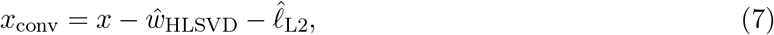

where 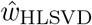 denotes the water component estimated using the rank-32 HLSVD procedure described above. The same L2 operator and field-strength-specific regularization settings were also used to construct the auxiliary input for the Y-Net models.

#### 2.4.2 L2-derived auxiliary input

For the Y-Net models, the subject-specific L2-derived lipid estimate described in Sec. 2.4.1 was used as an auxiliary network input. For a spectrum *x*, the original spectrum and its L2-derived lipid estimate were normalized using the same scaling factor *s* = max_*f*_ |*x*(*f*)|, yielding

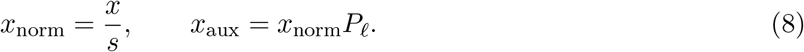

The auxiliary input therefore contained only the estimated lipid component, without HLSVD-based water removal. The normalized spectrum and its corresponding lipid estimate were processed by separate encoder branches of the Y-Net and combined in a shared decoder, as described below.

### 2.5 Network architecture and training

#### 2.5.1 Network architectures

All models employed one-dimensional convolutional encoder–decoder architectures to predict the combined water and lipid nuisance signal. Two architectural designs were considered: the single-encoder U-Net [41] and the dual-encoder Y-Net [32]. Complex-valued spectra were represented by separate real and imaginary channels.

##### Shared architectural details

Both architectures employed convolutional blocks consisting of two convolutions (kernel size 3) with PReLU activations. Downsampling was performed using max pooling, while upsampling used nearest-neighbor interpolation and skip connections from the corresponding encoder branches. Feature widths doubled at each downsampling level. Following the final decoder stage, features were additionally concatenated with the original input and initial convolutional features. A final 1 *×* 1 convolution predicted the real and imaginary components of the nuisance spectrum. Inputs were zero-padded to the next multiple of 2^*L*^, where *L* denotes the number of downsampling levels, and outputs were cropped to their original length.

Three network configurations were investigated:

##### Shallow Y-Net

As the starting point, we implemented a four-level Y-Net based on the architecture introduced in WALINET [32], retaining its dual-encoder structure and base filter multiplier of 16 (32 initial feature channels). The network processes the original spectrum and the L2-derived lipid estimate through separate encoders, with features from both branches combined at the bottleneck and through skip connections to a shared decoder.

##### Shallow U-Net

To investigate the contribution of the additional L2-derived input, a corresponding four-level U-Net was constructed using a single encoder. Its base filter multiplier was increased to 20 (40 initial feature channels) to compensate for the missing second encoder, enabling a comparison at equal depth and approximately matched capacity. The shallow U-Net and shallow Y-Net contained 6,589,872 and 6,867,036 trainable parameters, respectively.

##### Deep U-Net

The final WALRUS architecture used a higher-capacity U-Net with six downsampling levels and a base filter multiplier of 32 (64 initial feature channels), retaining the single-input design without an auxiliary prior. The network contained 268,565,664 trainable parameters.

#### 2.5.2 Network training and inference

All models were trained for 500 epochs using Adam to minimize the mean squared error between predicted and simulated complex nuisance spectra. The shallow networks used 285 batches of 2,000 spectra per epoch, while the deep U-Nets used 600 batches per epoch, with batch sizes of 950 at 7 T and 1,750 at 3 T. Batch sizes were adjusted according to GPU memory constraints. The initial learning rate was 2 *×* 10^*−*4^ and was halved after epochs 50, 100, 150, and 200.

The shallow networks were trained using the full native 7 T FID length of 840 samples. Acquisition-length augmentation was omitted to maintain matched training conditions for the U-Net/Y-Net comparison, as the L2 operator used for the Y-Net’s auxiliary input would need to be recomputed for each FID length. The deep U-Nets, which do not require an L2-derived auxiliary input, additionally employed acquisition-length augmentation, with FID lengths sampled uniformly between 420 and 840 samples at 7 T and between 144 and 288 samples at 3 T. This augmentation was introduced to accommodate different FID lengths across MRSI acquisition protocols, as truncating an FID is equivalent to shortening the acquisition while preserving the sampling interval. A single acquisition length was used for each training batch. Training and validation loss curves are shown in Supplementary Fig. S2.

#### *In vivo* inference

For inference, each *in vivo* spectrum *x* was normalized by *s* = max_*f*_ |*x*(*f*)| before being passed to the network. The predicted normalized nuisance signal *y* was rescaled by *s* and subtracted from the original spectrum, yielding *x*_clean_ = *x − sy*.

### 2.6 Experimental comparisons and evaluation

#### 2.6.1 Experimental comparisons

At 7 T, previous work demonstrated that simulation-based deep learning outperforms conventional HLSVD+L2 processing [32]. The present study therefore focused on further optimization of the simulation-based framework through controlled comparisons. At 3 T, where such improvements over conventional processing had not yet been established, WALRUS was evaluated directly against HLSVD+L2.

To assess the effect of simulator calibration, shallow U-Nets trained with heuristic and empirically calibrated parameter distributions were compared under otherwise identical conditions. The contribution of the L2-derived auxiliary input was subsequently evaluated by comparing calibrated shallow U-Nets with capacity-matched shallow Y-Nets. Each shallow-network configuration was trained with three independent simulation seeds. Due to its substantially higher computational cost, the final deep U-Net was trained using a single seed. To separately assess the contribution of on-the-fly simulation, an additional 7 T deep U-Net was trained using 1.2 million precomputed synthetic spectra generated at the full FID length of 840 samples. During training, spectra were randomly cropped for each batch to FID lengths between 420 and 840 samples, matching the acquisition-length augmentation of the on-the-fly model (Supplementary Fig. S2d).

For the 3 T evaluation, the deep U-Net was trained from scratch using a field-strength-specific spectral basis and independently calibrated simulation distributions.

#### 2.6.2 Evaluation of simulator realism in joint parameter space

Simulator realism was evaluated using parameter estimates from the five held-out 7 T UCSF datasets, without prior water or lipid removal. Each voxel was represented by relative metabolite amplitudes, SNR, frequency shift, and zero- and first-order phase. Water- and lipid-to-metabolite ratios were excluded because their estimation relied on a preliminary nuisance-removal model. FWHM was excluded because the LCModel-reported linewidth was not directly comparable to the simulator broadening parameter.

Each parameter *p* was robustly standardized using the median and IQR of the pooled *in vivo* values,

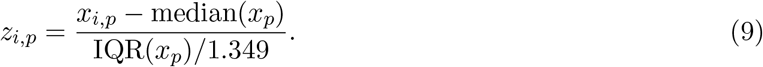

The same normalization was applied to simulated parameters. PCh was excluded because its *in vivo* IQR was zero.

For each distribution approach, 100,000 parameter vectors were generated. Similarity to the measured parameter space was quantified using the mean five-nearest-neighbour (5-NN) distance,

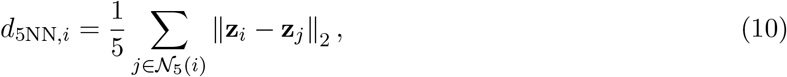

where *N*_5_(*i*) denotes the five nearest eligible *in vivo* neighbours. To establish an *in vivo* reference for the 5-NN distances, a leave-one-subject-out approach was used to avoid artificially small distances between spatially correlated voxels from the same subject. Simulated parameter vectors were compared against the pooled *in vivo* data from all five subjects. Lower distances indicate greater similarity to the observed parameter space.

#### 2.6.3 Simulation-based evaluation

Performance was evaluated using synthetic test spectra generated with the calibrated simulator: 100,000 spectra per held-out UCSF subject at 7 T and 30,000 spectra per held-out dataset at 3 T (12 datasets). Identical test spectra were used for all methods compared within each field strength.

Let *n*_*i*_ denote the ground-truth complex nuisance signal, 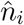 its estimate, and *m*_*i*_ the noise-free metabolite signal. Prediction errors were quantified using the nuisance- and metabolite-normalized residuals,

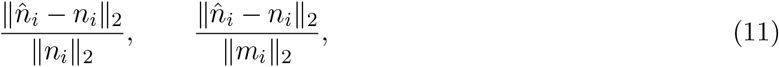

respectively, with the complex *l*_2_ norm evaluated over all spectral points.

Residuals were averaged across simulations within each subject or dataset. At 7 T, results from independently trained model seeds were additionally averaged within subjects, and mean *±* standard deviation (SD) was reported across the five subjects. At 3 T, mean *±* SD was reported across the 12 held-out datasets.

For conventional HLSVD+L2 processing, the water and lipid components were estimated independently from the complete simulated spectrum and summed to obtain 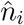 .

#### *2*.*6*.*4 In vivo* fit rates

*In vivo* performance was assessed using LCModel Cramér–Rao lower bounds (CRLBs) [42] for tNAA, tCr, tCho, and Glx. A voxel within the brain mask was considered successfully fitted if its CRLB was finite and satisfied 0 *<* CRLB *≤* 20%. The fit rate was defined as the fraction of brain-mask voxels meeting this criterion; all other voxels were counted as fitting failures.

Fit rates were calculated for the five held-out 7 T subjects and the 12 held-out 3 T datasets. At 7 T, values from independent model seeds were averaged within each subject. Results are reported as mean *±* SD across subjects or datasets, respectively.

#### 2.6.5 Sensitivity to the L2-derived auxiliary input

The sensitivity of the shallow Y-Net to its L2-derived auxiliary input was investigated using the three independently trained models and the five held-out 7 T UCSF subjects. Only the subject-specific L2 operator was modified; evaluation spectra and residual metrics remained unchanged.

##### Lipid-mask sensitivity

The extracranial lipid mask was eroded or dilated by one to four six-connected voxel layers. An L2 operator was recomputed for each mask using the original regularization target *τ* = 0.99. For dilated masks, newly included brain voxels contained the original spectrum after independent water removal, introducing metabolite contamination into the estimated lipid subspace.

##### Regularization sensitivity

Using the original lipid mask, the L2 operator was recalculated for trace targets *τ* = 0.91–1.00 in increments of 0.01, compared with the training target of *τ* = 0.99. At *τ* = 1, the lipid projection becomes zero and provides no auxiliary information.

For both sensitivity analyses, residuals were averaged across simulations and model seeds within each subject. Mean *±* SD was reported across the five subjects.

## 3 Results

### 3.1 Empirical calibration of simulation distributions improves 7 T performance

The empirically calibrated WALRUS sampling distributions more closely matched the measured *in vivo* parameter distributions than the heuristic distributions. Representative examples are shown in Fig. 2 a) and b) for relative NAA amplitude and frequency shift. Whereas the heuristic distributions provided broad generic coverage, the calibrated distributions were centered on the observed *in vivo* values while deliberately retaining extended tails beyond the measured range.

**Figure 2:**
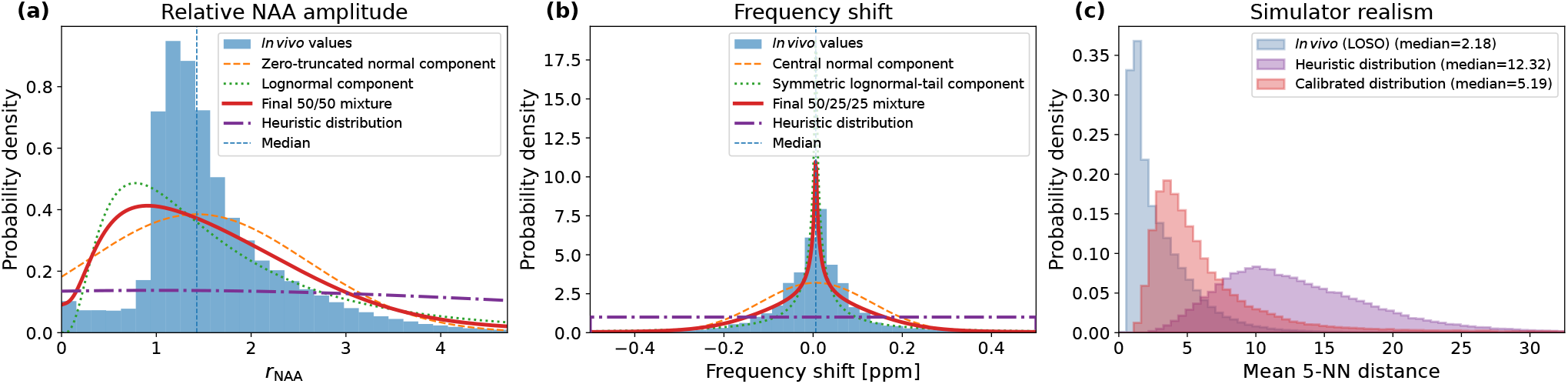
Empirical calibration and joint parameter-space realism of the WALRUS simulator. (a) Relative NAA amplitude and (b) frequency shift. Histograms represent the measured *in vivo* values, with the calibrated distribution and its mixture components overlaid together with the corresponding heuristic distribution and the *in vivo* median. (c) Distribution of the mean 5-nearest-neighbour (5-NN) distance in the joint parameter space for the leave-one-subject-out *in vivo* reference, calibrated simulations, and heuristic simulations. Lower distances indicate greater similarity to the measured *in vivo* parameter space. Distribution construction and calibration are described in Sec. 2.3.2 and Sec. 2.3.3; the joint parameter-space analysis is described in Sec. 2.6.2.

The improvement was also evident in the joint parameter-space analysis (Fig. 2c). The calibrated WALRUS simulations produced substantially smaller mean 5-NN distances to the measured *in vivo* parameter space than simulations generated using the heuristic distributions. The median mean 5-NN distance was 5.19 for calibrated and 12.32 for heuristic sampling, compared with 2.18 for the leave-one-subject-out *in vivo* reference. Thus, empirical calibration shifted the simulated parameter combinations substantially closer to those observed *in vivo*. At the same time, the calibrated distance distribution remained broader than the *in vivo* reference distribution, consistent with the deliberate broadening of the calibrated sampling distributions.

This improvement in simulation realism was also reflected in the quality of the *in vivo* metabolite maps (Fig. 3). The shallow U-Net trained with heuristic distributions and the shallow U-Net trained with empirically calibrated distributions used the same four-level architecture and training procedure and differed only in the parameter distributions used for simulation. The heuristic-training U-Net exhibited spatially localized fitting failures, including conspicuous gaps in the metabolite maps even in the superior slice, most prominently for tNAA. These failures were largely eliminated after training with the empirically calibrated distributions, resulting in substantially more spatially complete metabolite maps. The same pattern was observed across additional held-out UCSF subjects (Supplementary Fig. S1). The representative spectra from the corresponding locations showed the same trend (Supplementary Fig. S6), with the calibrated U-Net providing more consistent nuisance-signal removal than the heuristic-training U-Net.

**Figure 3:**
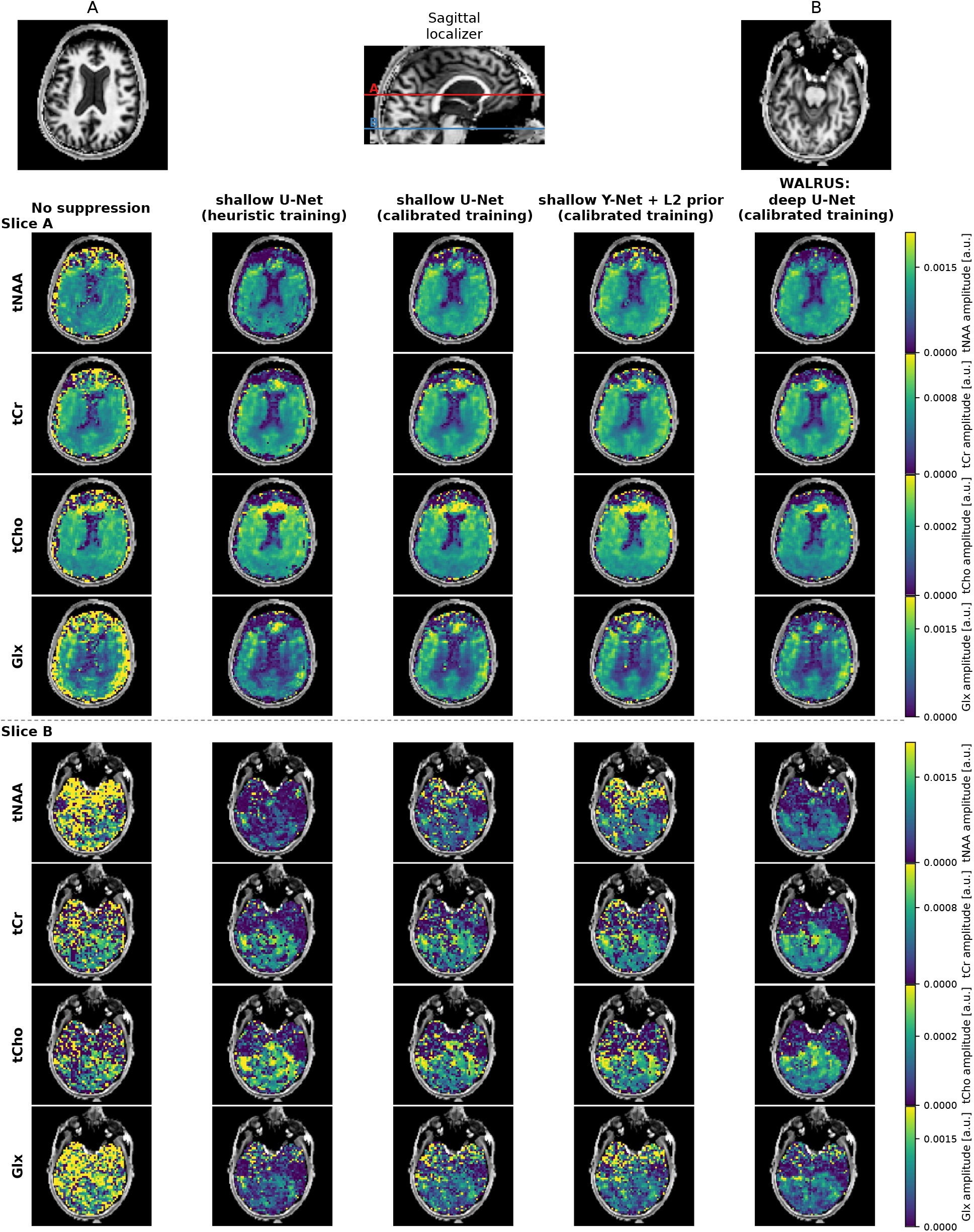
Qualitative 7 T comparison of WALRUS development stages. LCModel-derived tNAA, tCr, tCho, and Glx maps from a held-out UCSF subject are shown without nuisance removal and after applying four previously trained models: a shallow U-Net with heuristic or calibrated simulations, a capacity-matched Y-Net with L2 prior, and the final deep WALRUS U-Net. Superior (A) and inferior (B) slices are indicated on the sagittal localizer. Color scales are identical across methods and slices for each metabolite.

The qualitative improvement was consistent with the quantitative evaluation (Fig. 4a–c). Compared with the U-Net trained using heuristic distributions (orange), the U-Net trained using empirically calibrated distributions (yellow) showed lower nuisance-normalized and metabolite-normalized residuals and higher *in vivo* LCModel fit rates. Fit rates increased from 75.1% to 80.8% for tNAA, from 80.9% to 83.6% for tCr, from 82.3% to 83.3% for tCho, and from 65.5% to 74.1% for Glx. The corresponding learning curves further showed a markedly higher loss plateau for the heuristic U-Net than for the calibrated U-Net, both during training and when evaluated on validation spectra drawn from their respective simulation distributions (Supplementary Fig. S2a). Training and validation losses remained closely matched within each condition.

**Figure 4:**
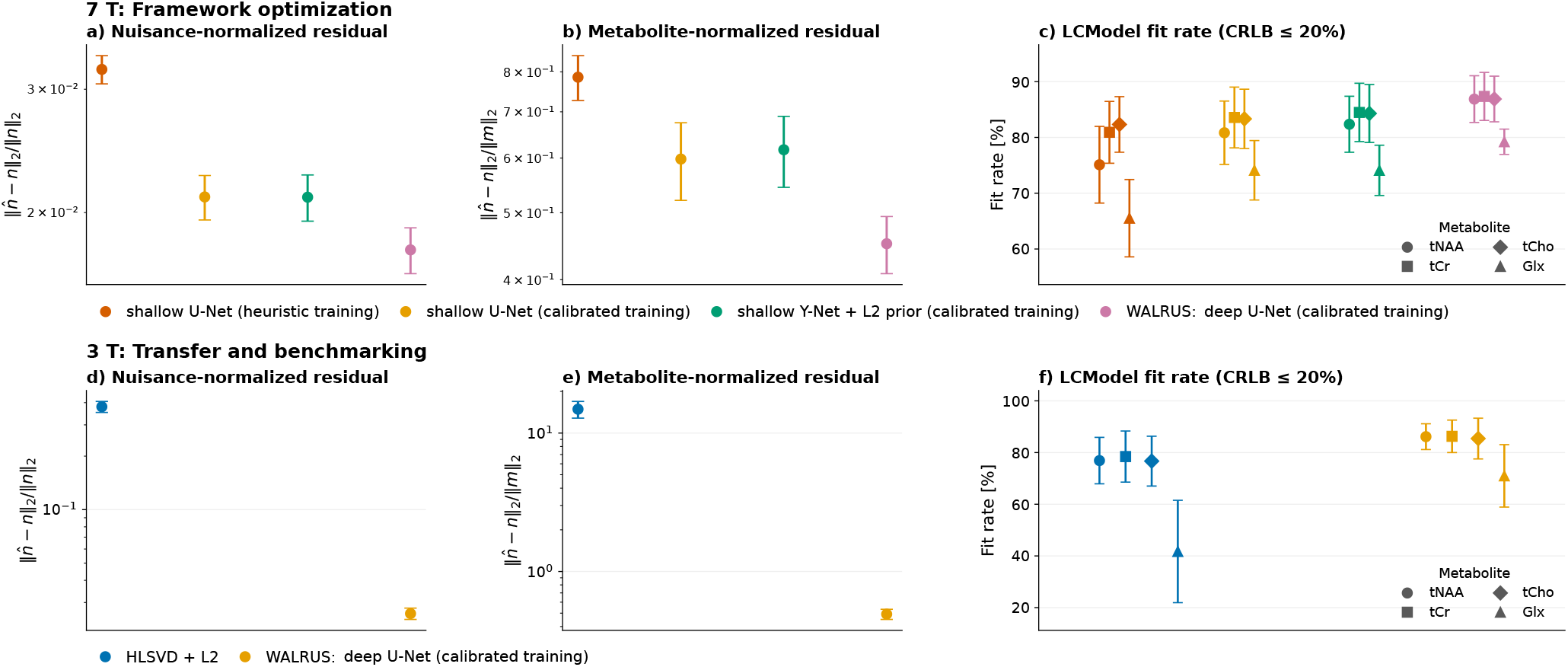
Quantitative evaluation of WALRUS at 7 T and 3 T. Top row (7 T): Framework optimization comparing the shallow U-Net trained with heuristic distributions, the shallow U-Net trained with calibrated distributions, the shallow Y-Net with L2-derived auxiliary input trained with calibrated distributions, and the final deep U-Net trained with calibrated distributions. **Bottom row (3 T):** Evaluation of the final deep U-Net against conventional HLSVD+L2 processing. Panels (a,d) show nuisance-normalized residuals and (b,e) metabolite-normalized residuals on synthetic test spectra. Panels (c,f) show *in vivo* LCModel fit rates (CRLB *≤* 20%) for tNAA, tCr, tCho, and Glx. Points and error bars indicate mean and sample standard deviation across five held-out 7 T subjects and twelve 3 T datasets, respectively. Results from multiple training seeds were averaged within each 7 T subject. Residuals are displayed on logarithmic scales. Residual errors are defined as 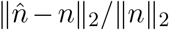 and 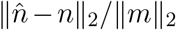, where 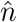 and *n* denote the estimated and ground-truth nuisance signals, respectively, and *m* is the noise-free metabolite signal.

### 3.2 Contribution and sensitivity of the L2-derived auxiliary input

The L2-derived auxiliary input provided no clear overall performance advantage over the prior-free U-Net. The calibrated U-Net (yellow in Fig. 4a–c) and the capacity-matched Y-Net with L2-derived auxiliary input (green) achieved similar simulation-based residual errors, with neither architecture consistently outperforming the other across the two metrics. Likewise, *in vivo* LCModel fit rates differed by less than 1.6 % between the two models for all evaluated metabolites.

The L2-derived input additionally introduced a dependence on the subject-specific L2 operator (Fig. 5). Altering the lipid-mask definition by progressive erosion or dilation resulted in modest changes in both residual metrics (Fig. 5 a,b). A stronger dependence was observed for the L2 trace target (Fig. 5c,d): performance varied as the trace target was moved away from the training value of 0.99 and deteriorated markedly at a target of 1.00, for which the lipid projection becomes zero and the auxiliary input contains no lipid information.

**Figure 5:**
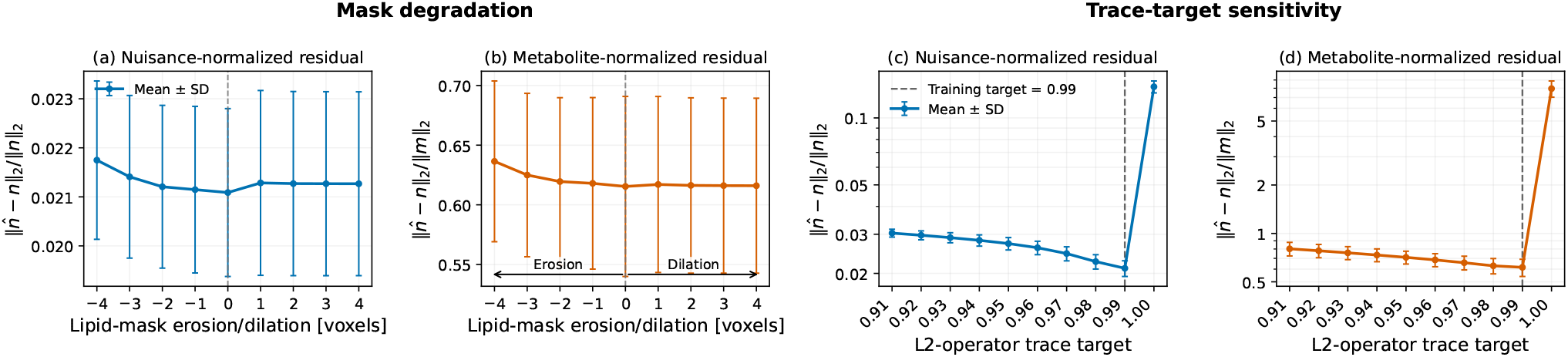
Sensitivity of the Y-Net to perturbations of the L2-derived auxiliary input. Left panels (a,b): Effect of lipid-mask erosion and dilation on the (a) nuisance-normalized and (b) metabolite-normalized residuals. Negative boundary shifts correspond to erosion of the lipid mask, whereas positive values correspond to dilation. At the strongest erosion (*−*4 voxels), 21.3 *±* 4.0% of the original lipid-mask voxels remained across the five UCSF subjects. **Right panels (c**,**d):** Effect of varying the L2-operator trace target on the (c) nuisance-normalized and (d) metabolite-normalized residuals. The Y-Net was trained using a trace target of 0.99 (dashed line); a target of 1.00 results in a zero L2 projection and therefore an uninformative auxiliary input. Values show mean *±* SD across the five held-out UCSF subjects after averaging across three independently trained model seeds.

### 3.2 Performance of the calibrated deep U-Net at 7 T

Building on the improvement achieved by empirical simulator calibration, the final higher-capacity WALRUS model provided a further qualitative improvement in the held-out 7 T data (Fig. 3). Compared with the calibrated shallow U-Net, the deep U-Net produced more spatially consistent metabolite maps, with the clearest additional improvements in the challenging inferior slice. The representative spectra showed a corresponding improvement in nuisance-signal removal across the selected locations (Supplementary Fig. S6). Overall, Fig. 3 illustrates the progression from heuristic to calibrated U-Net training and, subsequently, to the final deeper calibrated model. Exploratory experiments with a seven-level U-Net resulted in unstable training, and the six-level architecture was therefore retained as the final configuration.

This improvement was also reflected in the quantitative evaluation (Fig. 4a–c). The deep U-Net achieved the lowest nuisance-normalized and metabolite-normalized residual errors on the synthetic test spectra and the highest *in vivo* LCModel fit rates across all evaluated metabolites. Compared with the calibrated shallow U-Net, fit rates increased from 80.8% to 86.9% for tNAA, from 83.6% to 87.4% for tCr, from 83.3% to 86.9% for tCho, and from 74.1% to 79.2% for Glx.

For the final six-level 7 T architecture, on-the-fly simulation improved generalization to independently simulated validation spectra compared with training on a fixed precomputed dataset (Supplementary Fig. S2d). With all other training conditions and the validation set held identical, the on-the-fly model showed a smaller training–validation gap and reached a minimum validation MSE of 9.35 *×* 10^*−*6^, compared with 1.078 *×* 10^*−*5^ for the fixed-data model, corresponding to a 13.3% lower minimum validation error.

Inference runtime was evaluated on a single NVIDIA Tesla V100 GPU (32 GB) and an Intel Xeon E5-2698 v4 CPU. Processing one in vivo MRSI dataset required 45.2 s on the GPU (0.89 ms per spectrum) and 1364 s on the CPU (26.9 ms per spectrum).

Supplementary Fig. S5 additionally illustrates qualitatively consistent metabolite maps at shorter native acquisition lengths.

### 3.4 Transfer to 3 T

At 3 T, the independently recalibrated and retrained WALRUS framework consistently outperformed conventional HLSVD+L2 processing on the held-out test datasets.

Qualitatively, WALRUS provided more effective nuisance-signal removal than conventional HLSVD+L2 processing in the held-out 3 T data (Fig. 6). The HLSVD+L2-processed tNAA and Glx maps showed reduced signal intensity, whereas WALRUS better preserved the corresponding metabolite signal. In the representative spectra, residual water and lipid signals were also incompletely suppressed in several locations after HLSVD+L2 processing, while WALRUS achieved more complete nuisance removal with better preservation of the metabolite region.

**Figure 6:**
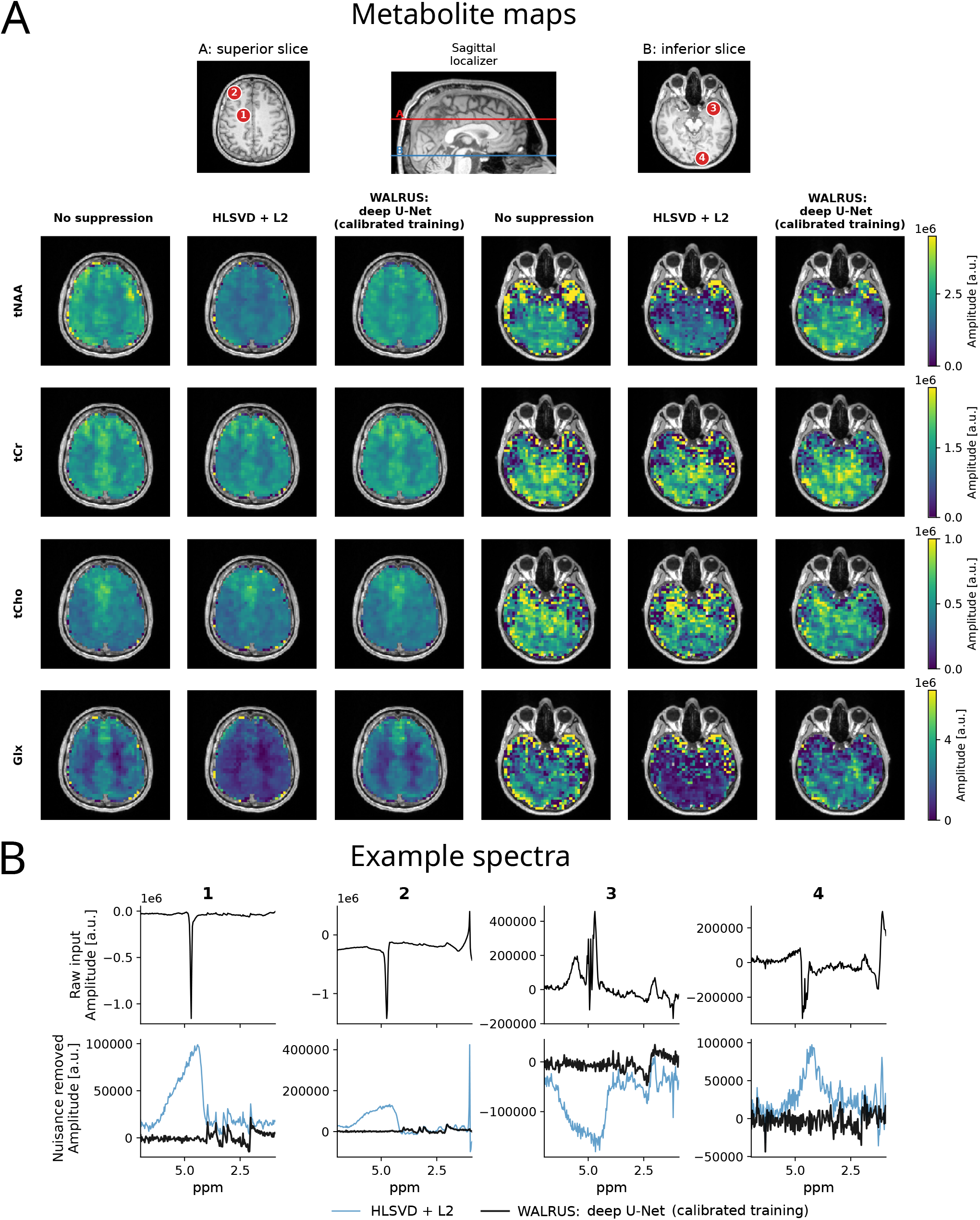
Qualitative 3 T comparison of conventional HLSVD+L2 and WALRUS. (A) LCModel-derived tNAA, tCr, tCho, and Glx maps from a held-out 3 T test dataset without nuisance-signal removal and after processing with conventional HLSVD+L2 and WALRUS. Superior and inferior slices are shown, with their locations indicated on the sagittal anatomical localizer. Identical color scaling was used across processing methods and slice locations for each metabolite. (B) Representative spectra from the four locations marked in (A), showing the raw input and the corresponding nuisance-removed spectra.

These qualitative findings were confirmed quantitatively across the 12 held-out 3 T datasets (Fig. 4d–f). On synthetic test spectra, WALRUS reduced the nuisance-normalized residual from 0.379 *±* 0.027 for HLSVD+L2 to 0.0260 *±* 0.0019, corresponding to a 93.2% reduction. The metabolite-normalized residual decreased from 14.88 *±* 2.05 to 0.492 *±* 0.041, corresponding to a 96.7% reduction.

The improvement was likewise reflected in the *in vivo* LCModel fit rates (Fig. 4f). Compared with HLSVD+L2, WALRUS increased the fit rate from 76.9% to 86.2% for tNAA, from 78.5% to 86.3% for tCr, from 76.7% to 85.4% for tCho, and from 41.7% to 71.0% for Glx. The largest absolute improvement was observed for Glx, with an increase of 29.3 percentage points.

## 4 Discussion

This work introduces WALRUS as a fitting-consistent, empirically calibrated, and prior-free framework for deep-learning-based water and lipid removal in ^1^H-MRSI.

### 4.1 Empirical calibration of the simulation space

The choice of simulation parameter distributions is a central consideration in supervised deep learning for MRS. WALINET demonstrated the effectiveness of combining simulated metabolite spectra with *in vivo*-derived water and lipid signals, using broad parameter distributions to cover diverse spectral conditions [32]. While this strategy enabled robust nuisance removal across different acquisition settings, the influence of the underlying parameter distributions on network performance was not systematically investigated.

Our controlled experiments demonstrate that replacing broad heuristic distributions with empirically calibrated distributions substantially improves nuisance removal under otherwise matched training conditions. The calibrated simulations occupied a joint parameter space considerably closer to the measured *in vivo* data, despite retaining deliberately broadened distributions and independent parameter sampling. This improvement was accompanied by lower residual errors, higher LCModel fit rates, and fewer localized fitting failures in held-out *in vivo* data.

These findings highlight the importance of balancing simulation realism and variability. Previous investigations of deep-learning-based MRS quantification have demonstrated that networks can develop substantial biases toward their training distributions, particularly under low-SNR conditions [43]. While insufficient variability may therefore limit generalization to previously unseen spectral conditions, our results demonstrate that excessively broad heuristic sampling can also compromise performance. Consistent with this interpretation, the heuristic U-Net exhibited substantially higher training and validation losses than the calibrated model, suggesting that its broader simulation space imposed a more challenging learning problem.

Rather than reproducing the measured *in vivo* distribution exactly, WALRUS uses independently sampled, empirically calibrated marginal distributions that are deliberately broadened beyond the observed variability. This maintains coverage of less common parameter combinations while concentrating training samples in spectrally relevant regions. Although the optimal degree of broadening and the potential benefit of explicitly modeling parameter correlations remain open questions, our results demonstrate that empirical calibration provides a substantial advantage over the investigated heuristic parameterization.

### 4.2 Role of the L2-derived prior and pipeline simplification

Classical L2-based lipid suppression exploits the assumption that extracranial lipid signals can be represented by a spectral subspace that is sufficiently distinguishable from metabolite signals [22]. However, overlap between these subspaces can lead to incomplete lipid removal or unintended metabolite signal attenuation. WALINET combined this conventional approach with deep learning by providing an L2-derived lipid estimate as an auxiliary network input [32]. This hybrid strategy allows the network to exploit both the measured spectrum and explicit subject-specific information about lipid contamination.

We investigated whether this additional information remains beneficial when the network is trained on a sufficiently representative spectral simulation space. Our capacity-matched comparison showed no clear overall performance advantage for the Y-Net with L2-derived auxiliary input over the prior-free U-Net. Nevertheless, the Y-Net exhibited a marked performance deterioration when the auxiliary lipid information was entirely removed, indicating that the network had learned to rely on this input despite not achieving superior performance. These findings suggest that nuisance-signal characteristics learned from calibrated spectral simulations can provide an effective alternative to explicitly constructed subject-specific lipid priors under the investigated conditions.

Beyond comparable performance, removing the auxiliary input substantially simplifies inference by eliminating the need for subject-specific lipid-mask generation, HLSVD processing [27], and L2 operator construction at deployment [44]. Importantly, HLSVD and lipid masks remain essential components of the WALRUS training-data preparation, where they are used to extract empirical water and lipid signals for spectral simulation. However, these steps are no longer required when applying the trained network to previously unseen subjects. Although explicit spatial priors may remain advantageous under other acquisition conditions, our results demonstrate that effective nuisance removal does not necessarily require their incorporation as additional network inputs.

### 4.3 Model capacity and on-the-fly simulation

Increasing network depth and capacity has enabled substantial performance improvements in deep learning [45]. However, highly expressive networks can also memorize finite training datasets, making training diversity an important consideration for generalization [46]. Data augmentation is a well-established strategy to mitigate overfitting and can act as an effective form of regularization [47]. In simulation-based learning, continuously generating new training examples offers a related advantage by exposing the network to new realizations of the underlying parameter space throughout optimization.

WALINET employed a large precomputed synthetic training dataset, combined with online phase augmentation and dropout [32]. WALRUS instead uses a GPU-optimized simulator to generate new spectral realizations continuously during training. To isolate the effect of on-the-fly simulation, we compared two otherwise identical deep U-Nets within the WALRUS framework, differing only in whether training spectra were precomputed or generated on the fly. Despite using 1.2 million precomputed spectra and identical batch-wise random FID-length augmentation between 420 and 840 samples, the fixed-data model exhibited a substantially larger training–validation gap than the on-the-fly model. Continuous simulation also achieved a lower minimum validation MSE. These findings suggest that repeatedly resampling the underlying spectral parameters acts as an effective form of regularization, improving generalization to independently simulated validation spectra.

The final six-level WALRUS U-Net additionally achieved lower residual errors and higher *in vivo* LCModel fit rates than the calibrated shallow U-Net, consistent with the potential benefits of increased model capacity when sufficient training diversity is available. Further increasing the architecture to seven levels resulted in unstable training under the investigated configuration, making the six-level U-Net a practical compromise between model capacity and training stability. Together, these findings suggest that combining higher-capacity networks with continuously generated synthetic training data is an effective strategy for improving the performance of the calibrated WALRUS framework.

### 4.4 Transferability and application to 3 T

Differences in field strength, acquisition protocols, and scanner characteristics present substantial challenges for the generalization of MRS methods [48]. For simulation-based deep learning, adaptation to new acquisition settings is particularly challenging because the spectral basis and simulation parameter distributions must also reflect the changed acquisition conditions. WALRUS addresses this challenge through a systematic calibration strategy. Metabolite spectra are generated directly from the acquisition-specific basis set used for subsequent spectral fitting, which is typically already available in MRS workflows. Given representative *in vivo* data, the remaining simulation distributions can be empirically recalibrated using the same procedure. Although new calibration data are still required, this approach substantially reduces the need for manual simulator parameterization and facilitates adaptation to different acquisition settings.

The successful application at 3 T demonstrates the feasibility of this strategy. Using an acquisition-specific basis set and recalibrated simulation distributions, WALRUS was retrained without modifying the underlying framework. The resulting model substantially outperformed conventional HLSVD+L2 processing, achieving more effective nuisance removal and higher *in vivo* LCModel fit rates.

Notably, HLSVD+L2 attenuated tNAA and Glx signals despite leaving substantial residual lipid contamination, illustrating the trade-off between lipid suppression and metabolite preservation previously reported for NAA in [32]. WALRUS better preserved these signals while providing more effective nuisance removal.

### 4.5 Limitations and outlook

Three limitations should be considered. First, robustness to different acquisition lengths was evaluated only qualitatively. A systematic quantitative investigation of this capability was beyond the scope of the present study.

Second, generalization to pathological conditions was not evaluated. Previous MRSI studies in glioma and multiple sclerosis patients highlight the clinical relevance of such applications [49]. Particularly in brain tumors, lipid contamination poses additional challenges, as pathological lipid signals may overlap with extracranial lipid artifacts [17, 50]. While the broadened and factorized simulation distributions were designed to promote generalizability, systematic evaluation in patient cohorts, including the preservation of disease-related spectral features, remains an important direction for future work.

Finally, a direct end-to-end comparison between WALINET and WALRUS was not performed. Such a comparison could not be faithfully reproduced within the WALRUS simulator, as the frameworks differ in their spectral bases, signal normalization, network training strategies, and partially incompatible simulation parameter definitions. A direct benchmark would therefore require adapting the original WALINET pipeline to the present data. Instead, this study employed controlled ablation experiments within a consistent simulation framework to demonstrate the performance gains associated with individual methodological improvements. While these results establish clear benefits of the proposed changes, the overall performance gain relative to the original WALINET pipeline remains to be quantified.

## 5 Conclusion

Building on WALINET, this study systematically investigated key design choices for deep-learning-based water and lipid removal in ^1^H-MRSI. Controlled experiments demonstrated that empirically calibrated simulation distributions substantially improve nuisance removal and downstream metabolite fitting, while the L2-derived auxiliary prior provides no clear additional benefit. The final higher-capacity U-Net achieved further performance improvements, and on-the-fly simulation reduced overfitting compared with training on a large fixed synthetic dataset.

WALRUS combines these advances into an optimized, prior-free framework that eliminates the need for subject-specific lipid segmentation and L2 processing during inference. Its successful application at both 7 T and 3 T demonstrates the adaptability of the proposed simulation and training strategy to different acquisition settings.

## CRediT authorship contribution statement

**Hauke Fischer:** Conceptualization, Methodology, Software, Validation, Formal analysis, Investigation, Data curation, Writing – original draft, Visualization, Writing – review & editing. **Stanislav Motyka:** Conceptualization, Supervision, Writing – review & editing. **Paul Weiser:** Resources. **Bernhard Strasser:** Resources, Writing – review & editing. **Lukas Hingerl:** Resources, Writing – review & editing. **Korbinian Eckstein:** Resources. **Korawig Chaknam:** Validation. **Aaron P. Osburg:** Writing –review & editing. **Anton Altmeyer:** Writing – review & editing. **Bernardo Campilho:** Writing – review & editing. **Ekaterina Sazonova:** Resources. **Yan Li:** Resources. **Jing Liu:** Resources. **Barbara Dymerska:** Resources. **Zeinab Eftekhari:** Resources. **Thomas B Shaw:** Resources, Writing –review & editing. **Markus Barth:** Resources. **Gilbert Hangel:** Resources. **Sumit Kaushik:** Writing – review & editing. **Lukas Haider:** Resources. **Wolfgang Bogner:** Conceptualization, Writing – review & editing, Resources, Project administration, Funding acquisition.

## Data availability

The WALRUS implementation presented in this study is publicly available at [51]. The *in vivo* data used in this study are available from the corresponding author upon reasonable request for non-commercial research purposes and subject to institutional ethics approval and data protection regulations.

## Declaration of Generative AI and AI-assisted technologies in the writing process

During the preparation of this work the authors used ChatGPT 5.6 in order to improve the readability and language of the manuscript. After using this tool/service, the authors reviewed and edited the content as needed and take full responsibility for the content of the published article.

## Acknowledgments

Funded by the European Union (ERC, GLUCO-SCAN, 101088351). Views and opinions expressed are however those of the authors only and do not necessarily reflect those of the European Union or the European Research Council Executive Agency. Neither the European Union nor the granting authority can be held responsible for them. This research was funded in whole or in part by the Austrian Science Fund (FWF) [10.55776/P34198, 10.55776/KLI1089, 10.55776/J4820]. For open access purposes, the author has applied a CC BY public copyright license to any author-accepted manuscript version arising from this submission. The financial support by the Austrian Federal Ministry for Digital and Economic Affairs and the National Foundation for Research, Technology and Development, and the Christian Doppler Research Association, is gratefully acknowledged. The Department of Imaging Neuroscience at University College London is supported by the Wellcome Trust [226793/Z/22/Z]. B.D. is partially funded by the Medical Research Council [UKRI226]. ZE is supported by MNDRA Postdoctoral Fellowship: 2025002586. Data acquisition at UCSF was supported by a UCSF Research Evaluation and Allocation Committee (REAC) Award and the George A. Clough Memorial Endowment Fund.

