## Supplementary Material for "WALRUS: Robust and Transferable Deep Learning for Water and Lipid Removal in ^1^H-MRSI"

#### Supplementary Note S1: Simulator and calibration details

##### S1.1 Spectral simulation model

Each simulated spectrum was constructed as the sum of four signal components: metabolites, residual water, lipids, and receiver noise,

$$x(f) = m(f) + w(f) + \ell(f) + n(f), \quad \text{(S1)}$$

where $m(f)$ denotes the metabolite signal, $w(f)$ the residual-water component, $\ell(f)$ the lipid component, and $n(f)$ complex receiver noise. The following sections describe the generation of each component and their subsequent combination into the final simulated spectrum.

##### Metabolite model

The metabolite model was derived from the same LCModel basis set used for spectral fitting. Before use in simulation, the artificial reference resonance near 0 ppm contained in the LCModel basis was removed from each basis component using rank-5 HLSVD (1). Components with resonance frequencies between −0.2 and 0.2 ppm were reconstructed and subtracted from the basis FIDs. The resulting cleaned basis functions $b_k(t)$ were used throughout the simulation.

Let $c_k \geq 0$ denote the independently sampled amplitude of metabolite component $k$. Before application of line-shape and phase distortions, the metabolite FID was

$$m_0(t) = \sum_{k=1}^{K} c_k b_k(t). \tag{S2}$$

For each simulated spectrum, a common zero-order phase $\phi_0$, frequency offset $\Delta f$, Gaussian FWHM $G$, and Lorentzian FWHM $L$ were applied to the complete metabolite mixture. The Gaussian and Lorentzian broadening terms together produce a Voigt-type line shape:

$$m_1(t) = m_0(t) \exp[i\,(\phi_0 + 2\pi\Delta f\, t)] \exp\left[-\pi L t - \frac{(\pi G t)^2}{4\ln 2}\right]. \tag{S3}$$

Here, $\Delta f$, $G$, and $L$ are expressed in Hz, and $\phi_0$ in radians. A first-order phase term was subsequently applied in the frequency domain,

$$m(f) = \mathcal{F}\{m_1(t)\} \exp(i\phi_1 f), \tag{S4}$$

where $f$ denotes frequency in Hz relative to the spectral center ($f = 0$) and $\phi_1$ is expressed in rad/Hz. The signs of the frequency- and phase-shift terms follow the Fourier-transform convention used by the simulator.

Rather than sampling the Gaussian and Lorentzian linewidths independently, a target Voigt FWHM $V$ was sampled first. A line-shape fraction $r \sim \mathcal{U}(0, 1)$ was then used to continuously span the range from pure Gaussian ($r = 0$) to pure Lorentzian ($r = 1$) broadening. Using the standard approximation for the Voigt FWHM (2),

$$V \approx 0.5346L + \sqrt{0.2166L^2 + G^2}, \tag{S5}$$

the maximum Lorentzian linewidth corresponding to $G = 0$ was defined as

$$L_{\max} = \frac{V}{0.5346 + \sqrt{0.2166}}. \tag{S6}$$

The individual Lorentzian and Gaussian linewidths were then obtained as

$$L = rL_{\max}, \tag{S7}$$

$$G = \sqrt{(V - 0.5346L)^2 - 0.2166L^2}. \tag{S8}$$

##### Empirical water and lipid components

Residual-water and lipid line shapes were sampled from subject-specific *in vivo* resource pools rather than generated analytically. Brain and extracranial lipid masks were derived from segmentations of the anatomical T1-weighted images. Residual water was estimated using the HLSVD procedure described in the main paper. Water estimates from voxels within the brain mask formed the subject-specific water resource pool. Lipid resources were obtained from voxels within the extracranial lipid mask after subtraction of the corresponding HLSVD-derived water component.

Subjects were sampled uniformly, followed by uniform sampling of resources within the selected subject. Water and lipid resources were drawn from the same subject for each simulated spectrum. One water spectrum $\widetilde{w}(f)$ and $J = 10$ lipid spectra $\widetilde{\ell}_j(f)$ were selected. The lipid spectra were combined using positive random weights,

$$\begin{aligned} u_j &\sim \mathcal{U}(0,1), \\ \alpha_j &= \frac{u_j}{\sum_{q=1}^{J} u_q}, \\ \widetilde{\ell}(f) &= \sum_{j=1}^{J} \alpha_j \widetilde{\ell}_j(f). \end{aligned} \tag{S9}$$

Because the stored water resources were frequency-centered, the global frequency offset $\Delta f$ sampled for the metabolite component was also applied to the water signal,

$$\widetilde{w}_{\Delta f}(f) = \mathcal{F}\{\mathcal{F}^{-1}\{\widetilde{w}(f)\} \exp(i2\pi\Delta f t)\}. \tag{S10}$$

In contrast, the lipid spectra retained their measured frequency distribution and were not globally shifted. The simulated phase terms $\phi_0$ and $\phi_1$ were not applied to either empirical component, such that water and lipids retained their measured complex phase.

The peak spectral magnitudes of the metabolite, water, and lipid components were defined as

$$A_m = \max_f |m(f)|, \qquad A_w = \max_f |\widetilde{w}_{\Delta f}(f)|, \qquad A_\ell = \max_f |\widetilde{\ell}(f)|, \tag{S11}$$

where the maxima were taken over the full spectral bandwidth. Sampled dimensionless amplitude ratios $r_w$ and $r_\ell$ were then used to obtain the final water and lipid components,

$$w(f) = r_w A_m \frac{\widetilde{w}_{\Delta f}(f)}{A_w}, \tag{S12}$$

$$\ell(f) = r_\ell A_m \frac{\widetilde{\ell}(f)}{A_\ell}. \tag{S13}$$

Consequently, $r_w$ and $r_\ell$ specify their peak amplitudes relative to the peak metabolite amplitude.

##### Receiver noise and variable acquisition length

In addition to the metabolite, water, and lipid components, unscaled complex white noise $n_0(f)$ was generated as

$$n_0(f) = \epsilon_\mathrm{R}(f) + i\,\epsilon_\mathrm{I}(f), \tag{S14}$$

where the real and imaginary components were sampled independently from $\mathcal{N}(0,1)$. At this stage, no SNR-dependent scaling was applied.

All four components were initially represented at the native in vivo FID length of the corresponding acquisition setting. For models using acquisition-length augmentation, an acquisition length $N_\mathrm{acq}$ was sampled for each training batch and applied to all spectra within that batch. The metabolite, water, lipid, and unscaled noise components were transformed separately to the time domain, truncated to the first $N_\mathrm{acq}$ samples, and subsequently transformed back to the frequency domain.

The resulting acquisition-length-adjusted components are denoted by $m_\mathrm{acq}(f)$, $w_\mathrm{acq}(f)$, $\ell_\mathrm{acq}(f)$, and $n_{0,\mathrm{acq}}(f)$, respectively. They remained separate for subsequent noise scaling and assembly of the final simulated spectrum.

**SNR-based noise scaling and signal normalization**

After adjustment to the sampled acquisition length, the noise amplitude was scaled to reproduce a sampled target SNR. The SNR was defined as

$$\mathrm{SNR} = \frac{A_m}{2\sigma_n}, \qquad A_m = \max_f |m_{\mathrm{acq}}(f)|, \tag{S15}$$

where $A_m$ denotes the peak magnitude of the acquisition-length-adjusted metabolite spectrum and $\sigma_n$ the amplitude of the real component of the final complex noise. For the unscaled noise realization $n_{0,\mathrm{acq}}(f)$, the corresponding noise amplitude was calculated as

$$\sigma_{n_0} = \sqrt{\frac{1}{N_{\mathrm{acq}}} \sum_{i=1}^{N_{\mathrm{acq}}} \left[\Re\left\{n_{0,\mathrm{acq}}(f_i)\right\}\right]^2}. \tag{S16}$$

For a sampled target SNR, the required final noise amplitude is

$$\sigma_n = \frac{A_m}{2\,\mathrm{SNR}}, \tag{S17}$$

and the final noise component was obtained as

$$n(f) = \frac{\sigma_n}{\sigma_{n_0}}\, n_{0,\mathrm{acq}}(f) = \frac{A_m}{2\,\mathrm{SNR}\,\sigma_{n_0}}\, n_{0,\mathrm{acq}}(f). \tag{S18}$$

The unnormalized simulated spectrum was then assembled as

$$x_{\mathrm{raw}}(f) = m_{\mathrm{acq}}(f) + w_{\mathrm{acq}}(f) + \ell_{\mathrm{acq}}(f) + n(f). \tag{S19}$$

A common scaling factor was calculated from the peak magnitude of the complete simulated spectrum,

$$s = \max_f |x_{\mathrm{raw}}(f)|. \tag{S20}$$

The network input was obtained by normalizing the complete spectrum with this factor,

$$x(f) = \frac{x_{\mathrm{raw}}(f)}{s}. \tag{S21}$$

The corresponding training target consisted exclusively of the water and lipid components and was normalized using the same scaling factor,

$$y(f) = \frac{w_{\mathrm{acq}}(f) + \ell_{\mathrm{acq}}(f)}{s}. \tag{S22}$$

Using the same normalization factor for input and target preserves the relative amplitudes of the nuisance components with respect to the complete simulated spectrum.

#### S1.2 Parameter calibration

**Directly calibrated parameters.** Parameters directly available from the LCModel output, including SNR, frequency shift, phase parameters, and linewidth, were calibrated from their pooled voxel-wise values as described in the main paper. A broadening factor of $\kappa = 2$ was applied to all calibrated parameter distributions, increasing the probability of sampling less frequent extreme values while leaving the empirically determined distribution centers unchanged.

The sole exception was the linewidth parameter, for which $\kappa = 1$ was used. LCModel reports the total fitted spectral FWHM, whereas the Voigt FWHM parameter of the simulator describes additional broadening applied to the metabolite basis functions. The two quantities are therefore not directly equivalent, and the LCModel FWHM was used as an empirical proxy for the simulator broadening parameter. Applying the additional factor of $\kappa = 2$ to this distribution resulted in excessive sampling of very broad spectral lines; therefore, no additional broadening was applied to the linewidth distribution.

SNR values were additionally restricted to SNR $\geq 1$, corresponding to the minimum value reported by LCModel.

**Relative metabolite amplitudes.** Metabolite amplitudes required an additional normalization because the LCModel coefficients are not directly transferable to the amplitude scale used by the simulator. For each voxel $v$, the complete metabolite FID was first reconstructed from the fitted LCModel coefficients,

$$s_{\mathrm{met},v}(t) = \sum_{m=1}^{M} c_{m,v}\, b_m(t), \tag{S23}$$

where $c_{m,v}$ denotes the fitted coefficient of metabolite $m$ and $b_m(t)$ the corresponding complex LCModel basis FID, adapted to the bandwidth and number of time points used for simulation.

To account for the effect of voxel-wise linewidth on the spectral peak amplitude, the reconstructed metabolite FID was additionally broadened using the FWHM reported by LCModel. After conversion of the FWHM from ppm to Hz,

$$\widetilde{s}_{\mathrm{met},v}(t) = s_{\mathrm{met},v}(t) \exp\left(-\pi L_v t\right), \tag{S24}$$

where $L_v$ denotes the voxel-wise LCModel FWHM in Hz. For this normalization step, the LCModel linewidth was applied as a Lorentzian broadening term rather than using the stochastic Voigt broadening employed during spectral simulation.

A common metabolite reference amplitude was then obtained from the maximum magnitude of the complete broadened metabolite spectrum,

$$A_{\mathrm{met},v} = \max_f \left|\mathcal{F}\left\{\widetilde{s}_{\mathrm{met},v}(t)\right\}\right|. \tag{S25}$$

The relative amplitude of each metabolite was defined as

$$r_{m,v} = \frac{c_{m,v}}{A_{\mathrm{met},v}}. \tag{S26}$$

Thus, all metabolite coefficients within a voxel were normalized to the same reference amplitude derived from the complete metabolite spectrum, rather than to the peak amplitude of the individual metabolite. Positive finite values of $r_{m,v}$ were pooled across the calibration data separately for each metabolite and used as the measurements $x_j$ in the positive distribution model described in the main paper. Each metabolite distribution was calibrated independently using $\kappa = 2$.

The resulting distributions therefore describe relative basis amplitudes with respect to the peak magnitude of the complete metabolite signal rather than absolute metabolite concentrations.

**Water and lipid amplitude ratios.** Water- and lipid-to-metabolite amplitude ratios were not directly available from the spectral fitting results and were therefore estimated using a two-stage procedure. First, a preliminary WALRUS model was trained using the empirically calibrated distributions described above for all available parameters, while water and lipid amplitudes were sampled from the heuristic distributions used for the corresponding ablation models described in the main paper. For this preliminary model, the maximum lipid-to-metabolite ratio was limited to 70. The network architecture was identical to that of the four-level U-Net used in the subsequent ablation experiments.

The preliminary model was applied to the in vivo calibration data to predict the combined water and lipid nuisance component, $\widehat{s}_{\text{nuis},v}(t)$. A metabolite-dominated signal estimate was obtained as

$$\widehat{s}_{\text{met},v}(t) = s_{\text{full},v}(t) - \widehat{s}_{\text{nuis},v}(t). \tag{S27}$$

Residual water was reconstructed independently using HLSVD, following the same procedure used to generate the empirical water resources for training. The lipid contribution was then estimated as

$$\widehat{s}_{\text{lipid},v}(t) = \widehat{s}_{\text{nuis},v}(t) - s_{\text{water},v}(t). \tag{S28}$$

After Fourier transformation, the metabolite and water reference amplitudes were defined from their maximum spectral magnitudes,

$$A_{\text{met},v} = \max_{f} \left| \widehat{S}_{\text{met},v}(f) \right|, \tag{S29}$$

$$A_{\text{water},v} = \max_{f} \left| S_{\text{water},v}(f) \right|. \tag{S30}$$

Because the lipid estimate depends directly on the HLSVD-derived water component, residual water could otherwise dominate its maximum magnitude. The lipid amplitude was therefore determined only below 4 ppm,

$$A_{\text{lipid},v} = \max_{\delta(f)<4\,\text{ppm}} \left| \widehat{S}_{\text{lipid},v}(f) \right|. \tag{S31}$$

The voxel-wise water- and lipid-to-metabolite ratios were then defined as

$$r_{\text{water},v} = \frac{A_{\text{water},v}}{A_{\text{met},v}}, \tag{S32}$$

$$r_{\text{lipid},v} = \frac{A_{\text{lipid},v}}{A_{\text{met},v}}. \tag{S33}$$

The resulting positive ratios were pooled across the calibration data and used to parameterize the positive mixture distributions described in the main paper, using $\kappa = 2$. These calibrated distributions subsequently replaced the heuristic water and lipid amplitude distributions for WALRUS training.

#### Supplementary Note S2: Additional dataset information

Detailed acquisition parameters for the 7-T and 3-T datasets are summarized in Supplementary Table S2, following the MRSinMRS reporting recommendations. The distribution of 3-T acquisition configurations across scanners is given in Supplementary Table S1.

Table S1: Composition of the 3-T data used for simulator construction and evaluation. Each entry represents one scanner–subject–resolution dataset. The held-out datasets were not included in the training pool.

| Scanner/site | Matrix size | All | Training | Held-out |
|---|---|---|---|---|
| ClimaX, Brisbane | $36 \times 36 \times 23$ | 5 | 4 | 1 |
| | $50 \times 50 \times 31$ | 5 | 4 | 1 |
| | *Subtotal* | 10 | 8 | 2 |
| PRISMAFIT, Vienna | $36 \times 36 \times 23$ | 5 | 4 | 1 |
| | $50 \times 50 \times 31$ | 5 | 4 | 1 |
| | $64 \times 64 \times 41$ | 5 | 4 | 1 |
| | *Subtotal* | 15 | 12 | 3 |
| PRISMA, Vienna | $36 \times 36 \times 23$ | 5 | 4 | 1 |
| | $50 \times 50 \times 31$ | 5 | 4 | 1 |
| | $64 \times 64 \times 41$ | 5 | 4 | 1 |
| | $64 \times 64 \times 47$ | 5 | 4 | 1 |
| | *Subtotal* | 20 | 16 | 4 |
| VIDA, Vienna | $36 \times 36 \times 23$ | 5 | 4 | 1 |
| | $50 \times 50 \times 31$ | 5 | 4 | 1 |
| | $64 \times 64 \times 41$ | 5 | 4 | 1 |
| | $64 \times 64 \times 47$ | 1 | 1 | 0 |
| | *Subtotal* | 16 | 13 | 3 |
| **Total** | | **61** | **49** | **12** |

Table S2: Acquisition parameters of the 7-T and 3-T MRSI datasets. Parameters are reported according to the MRSinMRS recommendations where available. Shared protocol parameters are displayed across all four site/scanner columns. The scanner-specific distribution of the 3-T acquisition configurations is given in Supplementary Table S1.

| **Parameter** | **Scanner/site 1** | **Scanner/site 2** | **Scanner/site 3** | **Scanner/site 4** |
|---|---|---|---|---|
| **7-T dataset** | | | | |
| Site | Vienna | Brisbane | London | UCSF |
| Healthy volunteers | 5 | 5 | 5 | 5 |
| MR system | Siemens MAGNETOM 7T DotPlus | Siemens MAGNETOM 7T DotPlus | Siemens MAGNETOM Terra | Siemens MAGNETOM 7T DotPlus |
| System software | VE12U AP01 | VE12U AP01 | VE12U SP01 | VE12U AP01 |
| RF coil | Nova Medical, 1Tx/32Rx | Nova Medical, 1Tx/32Rx | Nova Medical, 8Tx/32Rx; quadrature-like TrueForm mode | Nova Medical, 1Tx/32Rx |
| *MRSI acquisition* | | | | |
| Sequence | 3D concentric-ring-trajectory free-induction-decay MRSI (3D-CRT-FID-MRSI) | | | |
| Spatial matrix | $64 \times 64 \times 35$ | | | |
| Field of view [$mm^3$] | $220 \times 220 \times 120.2$ | | | |
| Nominal voxel size [$mm^3$] | $\approx 3.4 \times 3.4 \times 3.4$ | | | |
| TR [ms] | 450 | 420 | 420 | 420 |
| Acquisition time [min] | $\approx 13.1$ | $\approx 12.2$ | $\approx 12.2$ | $\approx 12.2$ |
| Acquisition delay [ms] | 1.3 | | | |
| Spectral bandwidth [Hz] | 2778 | | | |
| Readout | Concentric rings with variable temporal interleaves | | | |
| Water suppression | Optimized WET | | | |
| $B_0$ shimming | | DESS Advanced initially; GRE Brain or interactive shimming when needed | Siemens WIP1441B; brain-mask-based shim optimization | GRE Brain initially; frequently supplemented with DESS Advanced |
| *Anatomical acquisition* | | | | |
| Sequence | $T_1$-weighted MP2RAGE | | | |
| Voxel size [$mm^3$] | $0.8 \times 0.8 \times 0.8$ | | | |
| Spatial matrix | $192 \times 224 \times 256$ | | | |
| TR / TE [ms] | 4300 / 2.4 | | | |
| Acquisition time [min:s] | 6:54 | | | |
| **3-T dataset** | | | | |

*Continued on next page*

*Supplementary Table S2 (continued)*

| **Parameter** | **Scanner/site 1** | **Scanner/site 2** | **Scanner/site 3** | **Scanner/site 4** |
|---|---|---|---|---|
| Site | Brisbane | Vienna | Vienna | Vienna |
| Healthy volunteers | 5 | 5 | 5 | 5 |
| MRSI acquisitions | 10 | 20 | 15 | 16 |
| MR system | Siemens MAGNETOM ClimaX | Siemens MAGNETOM Prisma | Siemens MAGNETOM PrismaFit | Siemens MAGNETOM Vida |
| System software | XA60 | XA60 | VE11C | XA60 |
| RF coil | Siemens 64-channel receive-only head coil | | | |
| *MRSI acquisition* | | | | |
| Sequence | 3D concentric-ring-trajectory free-induction-decay MRSI (3D-CRT-FID-MRSI) | | | |
| Spatial matrices | $36 \times 36 \times 23$, $50 \times 50 \times 31$, $64 \times 64 \times 41$, and $64 \times 64 \times 47$; scanner-specific allocation in Supplementary Table S1 | | | |
| Nominal voxel sizes [$mm^3$] | $\approx 6.1 \times 6.1 \times 6.1$, $4.4 \times 4.4 \times 4.5$, and $3.4 \times 3.4 \times 3.4$ (two superior–inferior coverages for the 3.4-mm protocol) | | | |
| In-plane FOV [$mm^2$] | $220 \times 220$ | | | |
| Through-plane FOV [mm] | 140 for the 6.1-mm, 4.4-mm, and standard 3.4-mm protocols; 160 for the extended-coverage 3.4-mm protocol | | | |
| TR [ms] | 450 | 450 | 450 | 455 |
| Acquisition time [min] | $\approx 2.5$–9.2, depending on spatial resolution and coverage | | | |
| Acquisition delay [ms] | 1.0 | | | |
| Spectral bandwidth [Hz] | $\approx 940$ | | | |
| Water suppression | WET | | | |
| *Anatomical acquisition* | | | | |
| Sequence | $T_1$-weighted MP2RAGE | | | |
| Voxel size [$mm^3$] | $0.9 \times 0.9 \times 0.9$ | | | |
| Spatial matrix | $176 \times 240 \times 256$ | | | |
| TR / TE [ms] | 1900 / 2.3 | | | |
| Acquisition time [min:s] | 4:10 | | | |

The 3-T dataset comprised 61 MRSI acquisitions obtained across four scanners and multiple spatial resolutions. Four subjects per scanner were assigned to training and one subject per scanner was held out for evaluation. Because several subjects were acquired at multiple spatial resolutions, this resulted in 49 training and 12 held-out datasets.

The scanner- and resolution-specific distribution of the 3-T data is summarized in Table S1.

### Supplementary Note S3: Additional analyses

#### S3.1 Additional qualitative evaluation of simulator calibration

To assess whether the qualitative effect of simulator calibration was consistent across held-out subjects, metabolite maps were additionally compared in three UCSF test subjects (Fig. S1). The two U-Nets used the same network architecture and training configuration and differed only in the parameter distributions used for synthetic training-data generation. Across subjects, the U-Net trained with heuristic WALINET-style distributions showed more spatially localized fitting failures, whereas training with the empirically calibrated WALRUS distributions resulted in more spatially complete metabolite maps.

#### S3.2 Training behavior

Training and validation loss curves for the calibrated four-level U-Net, the calibrated Y-Net with L2-derived auxiliary input, and the final WALRUS models at 3 T and 7 T are shown in Fig. S2. Losses decreased throughout training and approached stable values for all model configurations. For the final 7-T architecture, the otherwise matched comparison between on-the-fly simulation and a fixed precomputed training set additionally showed a smaller training–validation gap for on-the-fly training. All models were trained for the full 500 epochs.

#### S3.3 Representative simulator output

Representative spectra generated by the WALRUS simulator at 3 T and 7 T are shown in Fig. S3. The examples illustrate the range of simulated metabolite, residual-water, lipid, and noise contributions. For each example, the complete simulated spectrum is shown together with the corresponding combined nuisance component and metabolite component.

#### S3.4 L2 regularization parameter selection

The field-strength-specific L2 trace targets used throughout the study were chosen to balance lipid suppression against preservation of metabolite signal. Fig. S4 illustrates the effect of the trace target on the estimated lipid component at 3 T and 7 T. Lower trace targets resulted in stronger lipid projection, whereas higher trace targets produced progressively weaker lipid estimates. Based on this trade-off, trace targets of 0.97 at 3 T and 0.99 at 7 T were used throughout the study.

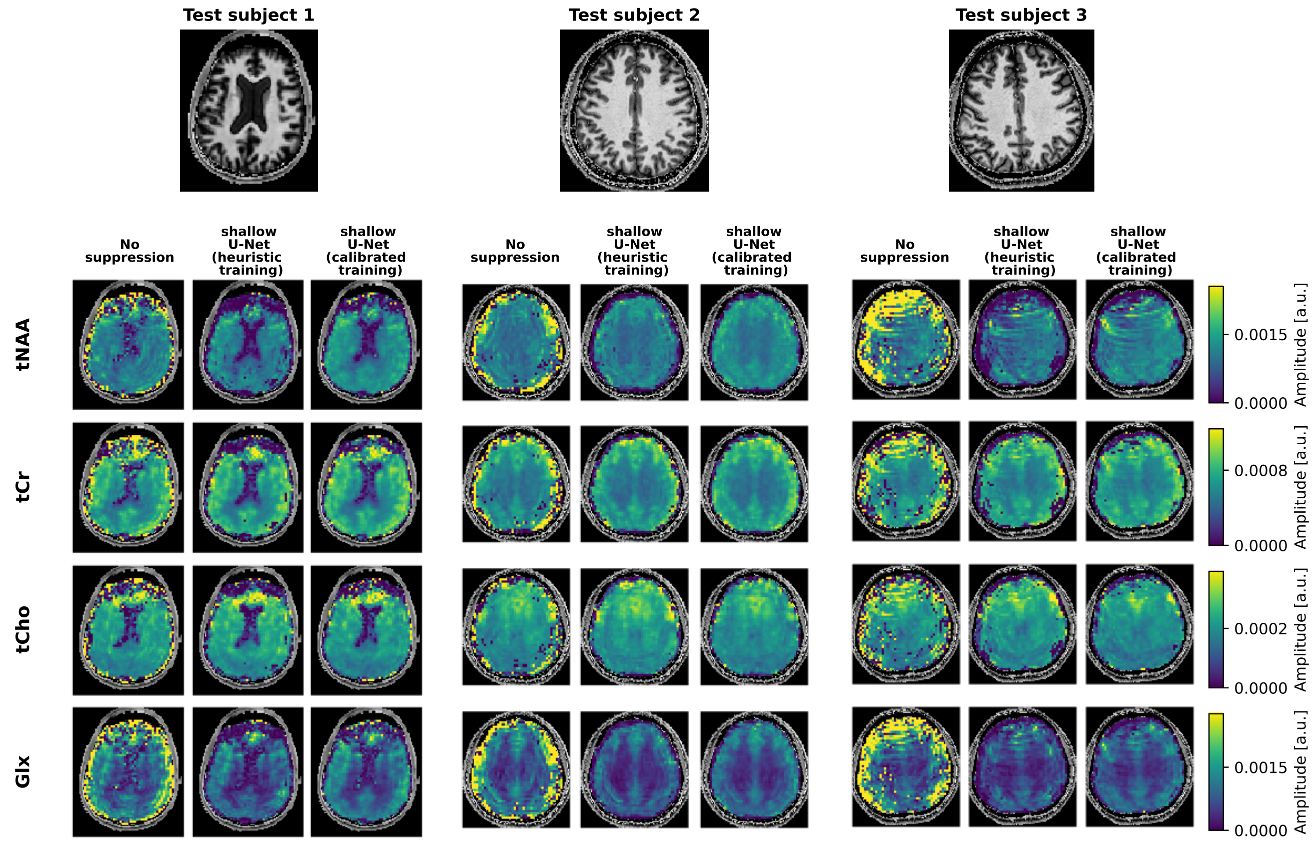


Figure S1: **Qualitative effect of empirical simulator calibration at 7 T.** LCModel-derived metabolite maps from three held-out UCSF test subjects are shown for spectra without nuisance-signal removal, after processing with a U-Net trained using the heuristic WALINET-style simulation distributions, and after processing with the same U-Net architecture trained using the empirically calibrated WALRUS distributions. The two U-Net conditions used the same architecture and training configuration and differed only in the parameterization of the synthetic training data. Maps are shown for total NAA (tNAA = NAA + NAAG), total creatine (tCr = Cr + PCr), total choline (tCho = GPC + PCh), and Glx (Glu + Gln). For each metabolite, identical color scaling was used across subjects and processing methods.

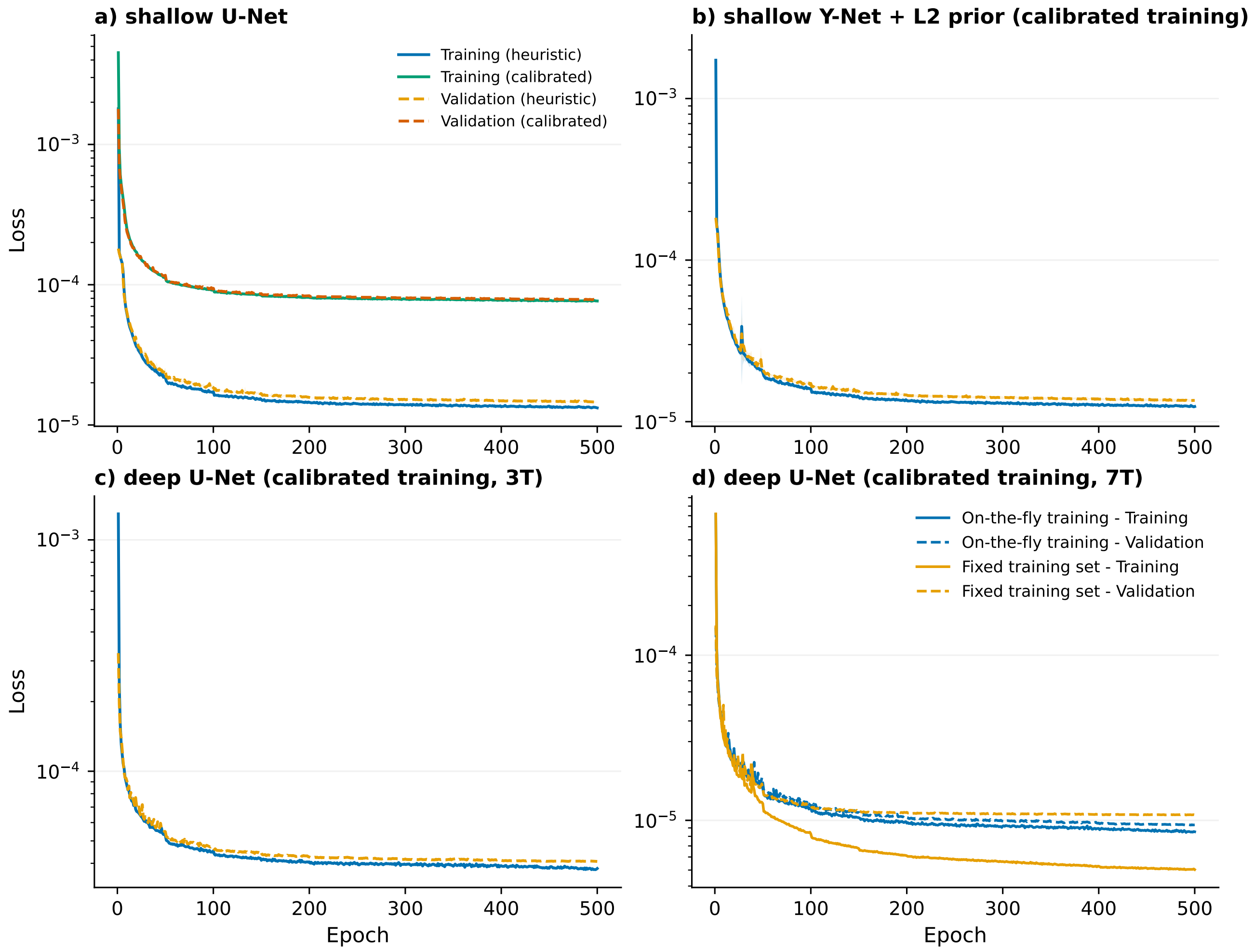


Figure S2: **Training and validation loss curves.** Training and validation losses over 500 epochs are shown for (a) the calibrated U-Net, (b) the calibrated Y-Net with L2-derived auxiliary input, and the final WALRUS models at (c) 3 T and (d) 7 T. Panel (d) additionally compares the final 7-T architecture trained using on-the-fly simulation with an otherwise identically trained model using a fixed precomputed synthetic training set. All models were trained for the full 500 epochs, and the models used for evaluation correspond to the final epoch rather than a validation-selected checkpoint.

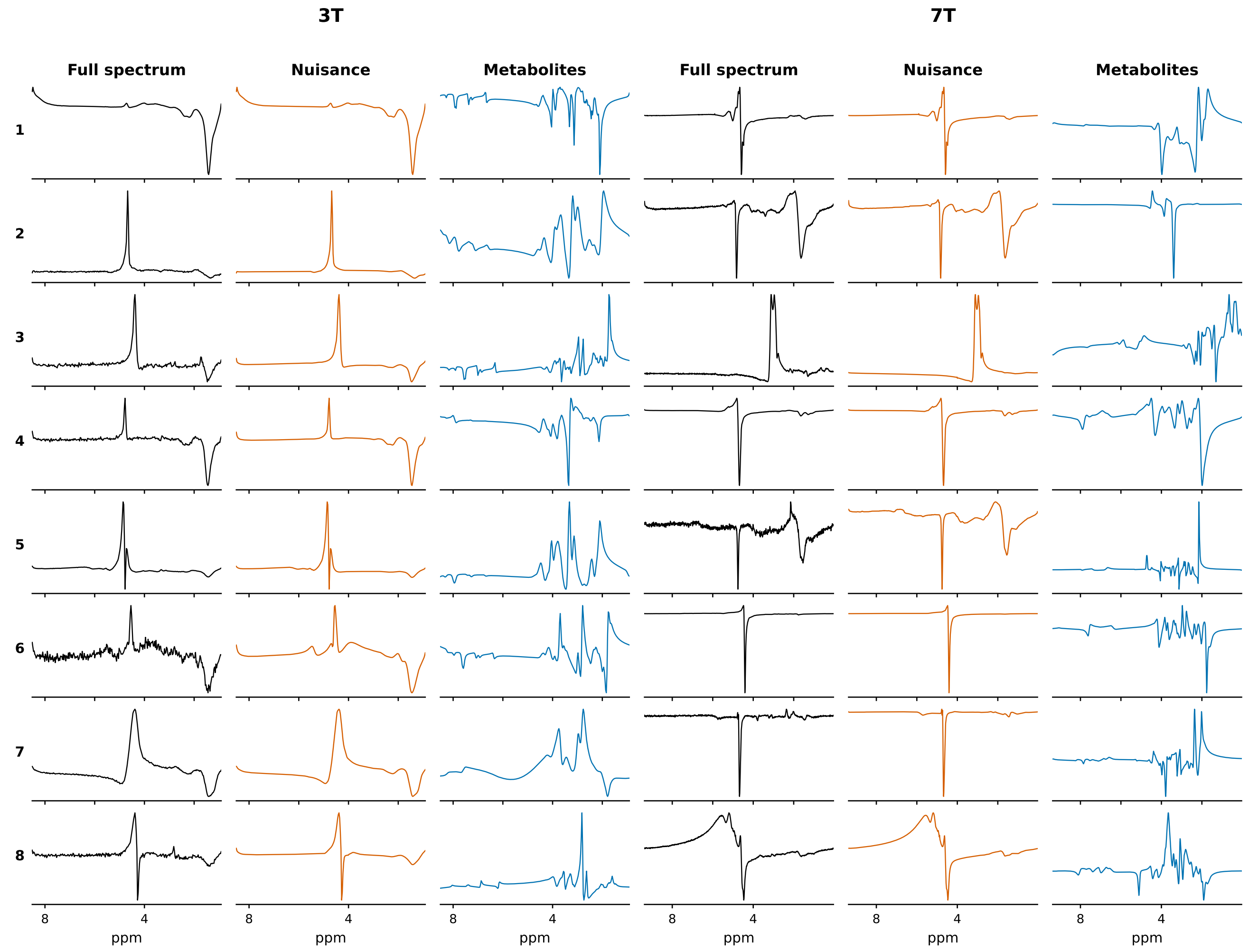


Figure S3: **Examples of synthetic spectra generated by the WALRUS simulator.** Eight representative examples are shown for 3 T (left) and 7 T (right). For each example, the real parts of the complete simulated spectrum, the combined water-and-lipid nuisance component, and the metabolite component are shown separately.

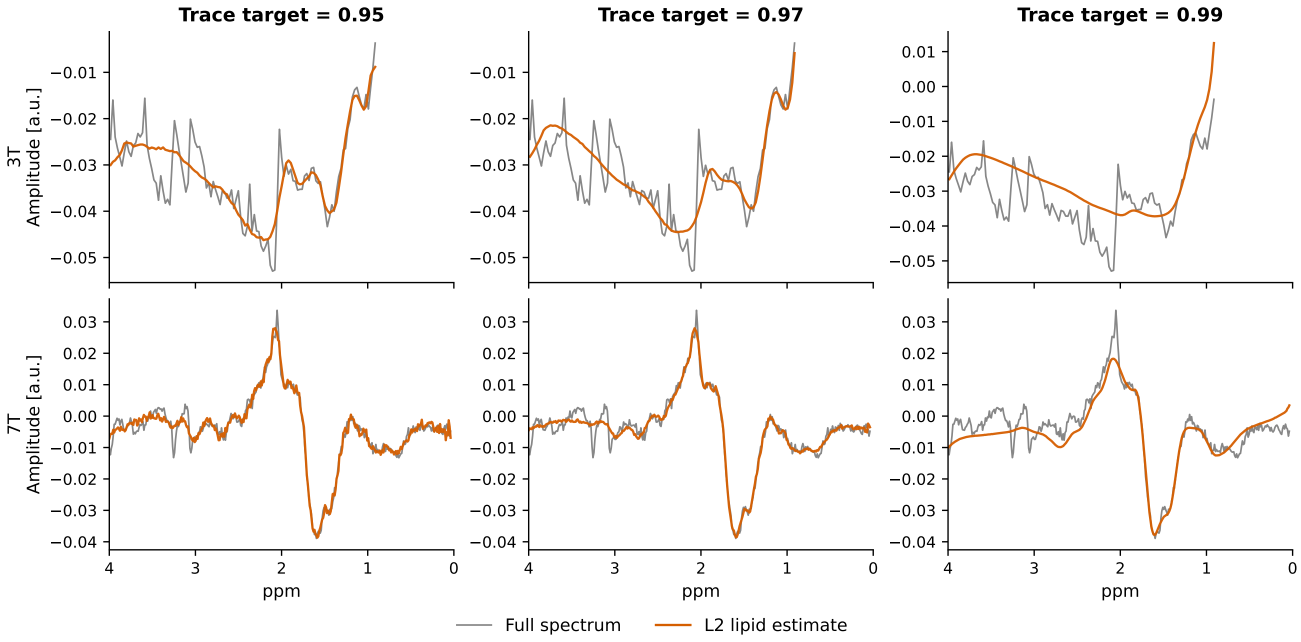


Figure S4: **Effect of the L2-operator trace target on the estimated lipid component.** Representative examples are shown at 3 T (top) and 7 T (bottom) for trace targets of 0.95, 0.97, and 0.99. The full spectrum is shown in gray and the corresponding L2-derived lipid estimate in orange.

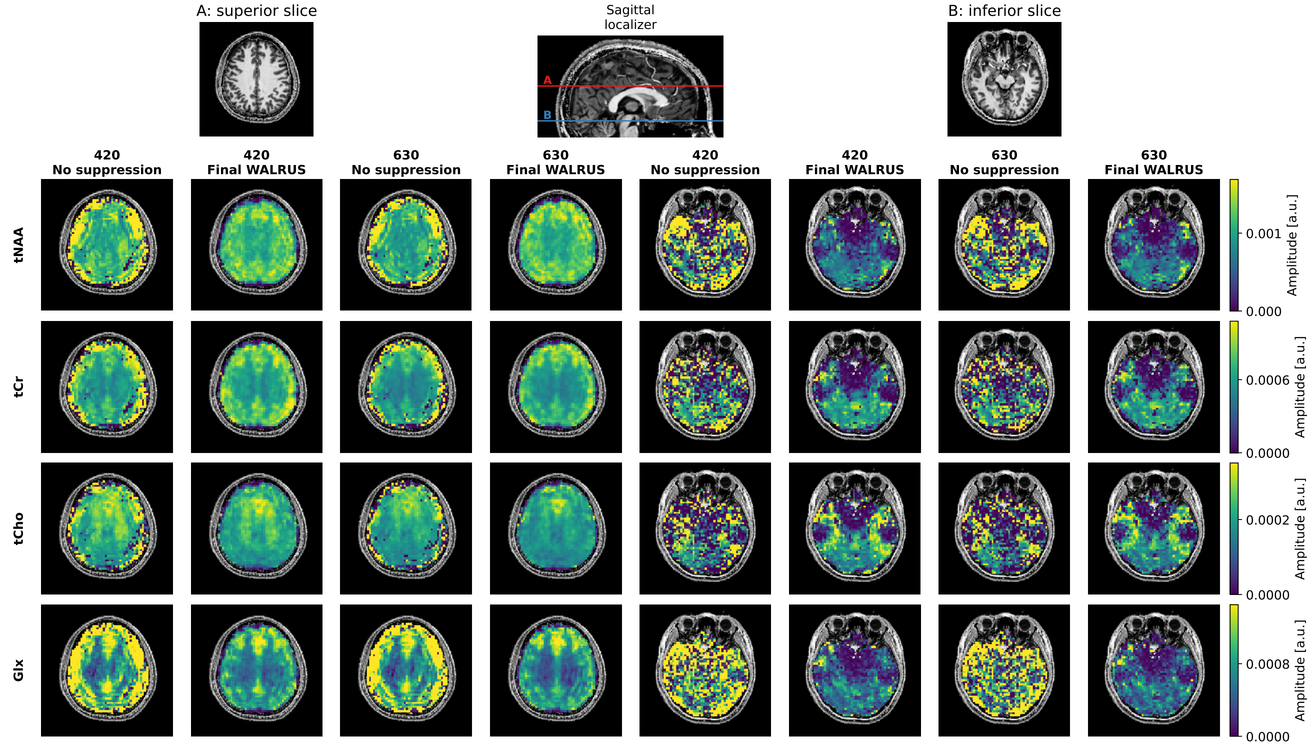


Figure S5: **Qualitative robustness to acquisition length.** Representative metabolite maps from one held-out 7-T subject after retrospective truncation to 420 and 630 acquired FID points. For each acquisition length, maps are shown before nuisance-signal removal and after processing with Final WALRUS (deep U-Net with calibrated training) for a superior (A) and an inferior (B) slice. tNAA, tCr, tCho, and Glx maps remain qualitatively consistent across both acquisition lengths, including the more challenging inferior slice. The native 840-sample acquisition corresponds to the standard evaluation setting used throughout the main paper and is therefore not repeated here.

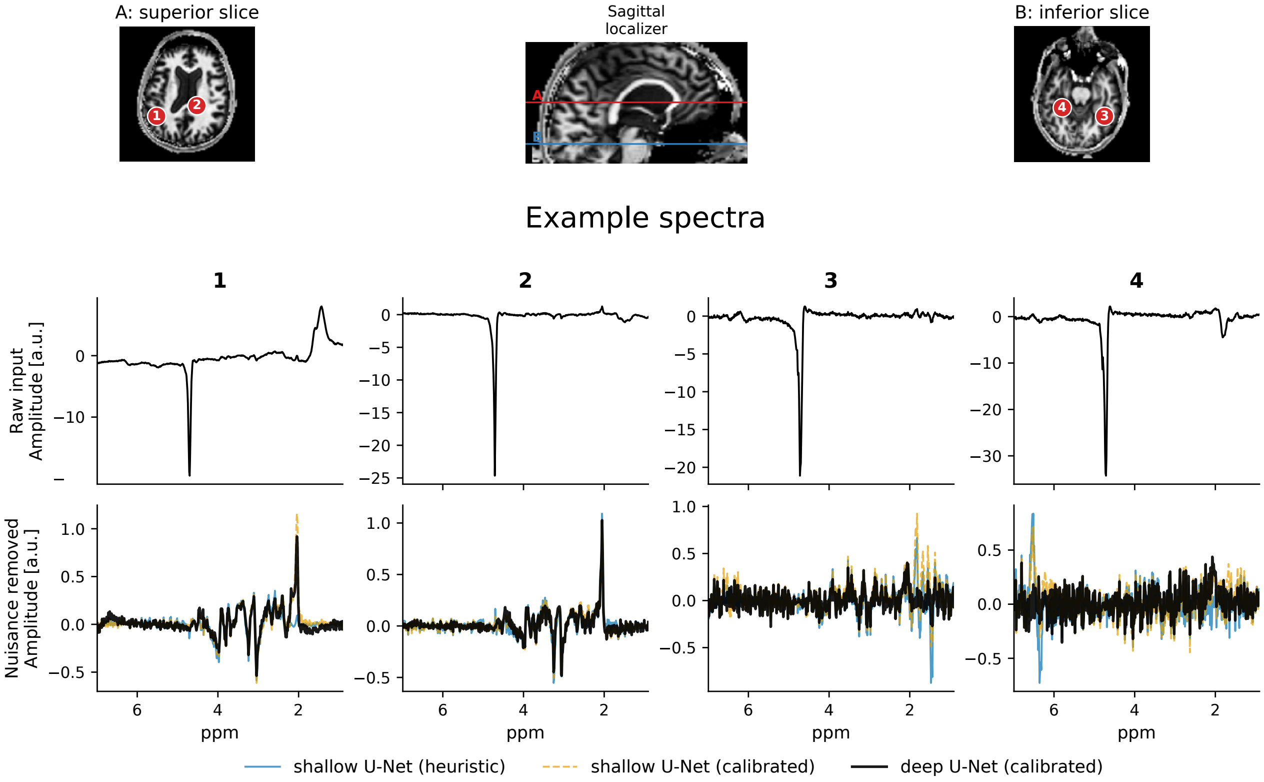


Figure S6: **Example spectra at 7T for different methods.**
